# *HTRA1-AS1*, an *ARMS2*-region long non-coding RNA, is downregulated in retinas of age-related macular degeneration patients

**DOI:** 10.1101/2025.10.29.25338834

**Authors:** Ping-Wu Zhang, Zi-He Wan, Sheng Liu, Jie Wang, Srinivasa Sripathi, Junyeop Ahn, Weifeng Li, Sean Li, Laura Fan, Cynthia A. Berlinicke, Jiang Qian, Shannath L. Merbs, Donald J. Zack

## Abstract

**Purpose:** The human 10q26 locus is a major genetic risk factor for age-related macular degeneration (AMD). Fine mapping by linkage and large-scale genome-wide association studies (GWAS) has narrowed this region to a 30-kb interval encompassing the *ARMS2* and *HTRA1* genes. However, the causative gene(s), risk variants, and underlying pathogenic mechanisms remain unresolved.

**Methods:** Long non-coding RNA (lncRNA) candidates within the *ARMS2–HTRA1* region were identified using human postmortem retinal RNA-seq data and public databases (NCBI, Ensembl). Candidate transcripts were validated by RT-PCR and Sanger sequencing. Published single-cell RNA-seq datasets were analysed to define cell type–specific expression, and RNA levels were compared between AMD and non-AMD donor retinas. Additionally, expression changes were assessed in human iPSC-derived retinal pigment epithelium (RPE) cells exposed to cigarette smoke extract (CSE) and paraquat (PQT).

**Results:** We identified and characterized the long noncoding RNA (lncRNA) HTRA1-AS1 and its transcript variants within the ARMS2 locus. HTRA1-AS1 overlaps ARMS2 and is transcribed in the antisense orientation. It is predominantly expressed in rod photoreceptors, Müller glia, and choroid/RPE tissues. Retinal HTRA1-AS1 expression was significantly reduced in AMD donors compared with controls (43 AMD vs. 44 controls, P = 0.007), whereas HTRA1 mRNA expression showed no significant difference (P = 0.121). In induced pluripotent stem cell (iPSC)-derived RPE cells, cigarette smoke extract (CSE)-induced oxidative stress markedly increased HTRA1-AS1 and ARMS2 expression by up to 8-fold and 75-fold, respectively, whereas paraquat treatment produced no significant changes.

**Conclusion:** These findings identify HTRA1-AS1 as an AMD-associated lncRNA within the ARMS2 locus that exhibits cell-type-specific expression, disease-associated downregulation, and responsiveness to cigarette smoke-induced oxidative stress. These results suggest that HTRA1-AS1 may contribute to AMD pathogenesis through noncoding regulatory mechanisms, warranting further functional investigation.

## Introduction

Age-related macular degeneration (AMD), the leading cause of irreversible blindness in the elderly, is characterized by the progressive degeneration of photoreceptors and retinal pigment epithelial (RPE) cells. ^1^ In 2019, the prevalence of early-stage AMD among adults aged 40 years and older in the United States was estimated at 11.64% .^2^ While aging, smoking, and diet constitute key environmental factors, numerous studies have established the importance of genetic contributions to AMD risk .^3^

Although approximately 70–80% of AMD cases are sporadic, genetic factors have been suggested to account for up to 70% of overall disease risk. ^4–5^ This contribution is comparable to that observed in Alzheimer’s disease (AD) and significantly greater than in other common neurodegenerative disorders like amyotrophic lateral sclerosis (ALS) and Parkinson’s disease (PD). For comparison, 90–95% of ALS and PD cases are sporadic, and known pathogenic variants explain only 10–20% of ALS and ∼14% of PD susceptibility. ^6–10^ Conversely, in late-onset AD, APOE ɛ4 is the strongest risk factor, and genome-wide association study (GWAS) identified variants collectively explain ∼70% of disease susceptibility. ^10, 11^

AMD risk factors are complex and multigenetic. To date, more than 40 genes and loci have been associated with AMD, with the complement factor H (*CFH*) locus on chromosome 1q31 and the *ARMS2/HTRA1* locus on chromosome 10q26 showing the strongest associations. ^5, 12,13^ Together, these two loci account for more than 50% of the inherited risk for AMD. ^14^ The remaining genetic risk is distributed across a variety of more minor loci that together account for about 20% of heritability. ^4, 5^ Ethnic variability has been observed for *CFH* variants, which show strong associations in Caucasian populations but inconsistent replication in some east Asian cohorts. ^15–17^ In contrast, the *ARMS2/HTRA1* locus has demonstrated robust associations across diverse populations, with no clear inter-ethnic differences. Furthermore, rod-mediated dark adaptation (RMDA) - a functional biomarker of early AMD - has been consistently linked to *ARMS2*, but not *CFH* risk alleles, even in individuals without overt disease. ^18^

At the functional level, the causal variation at the *CFH* locus is relatively well defined, implicating its role as a central regulator of the complement pathway. By contrast, the precise biological contribution of the *ARMS2/HTRA1* locus to AMD remains incompletely understood. *HTRA1* encodes a serine protease, but the function of *ARMS2* is still unclear, and the specific gene(s), risk variants, and molecular mechanisms driving AMD susceptibility at 10q26 are not yet established. ^5, 13, 19^

Given that the *ARMS2/HTRA1* locus remains one of the most strongly associated yet mechanistically least understood regions in AMD genetics, we sought to perform an in-depth investigation of this genetic interval. Our study aimed to identify novel coding or noncoding transcripts within the *ARMS2/HTRA1* region that may contribute to AMD pathogenesis and provide mechanistic insight into how this locus confers risk.

### Ethics statement

All aspects of this study were conducted in accordance with the principles of the Declaration of Helsinki, with informed consent being obtained from all participants. This project was approved by the Johns Hopkins University School of Medicine Institutional Review Board (IRB).

## Material and Methods

### Human AMD eye samples

Post-mortem eye tissues (AMD and non-AMD controls) were obtained from the National Disease Research Interchange (NDRI, Philadelphia, USA) and transported on wet ice. Processing of all samples occurred within 48 hours of death. The specific clinical staging of AMD donors were not provided by the National Disease Research Interchange (NDRI). Before retina isolation, macular photography was performed to document pathology. AMD samples were included only if they had documented clinical history and gross macular pathology consistent with disease. Tissue acquisition, handling, and processing were conducted in accordance with the ARVO Best Practices for Using Human Eye Tissue in Research (November 2021) and applicable Johns Hopkins University Institutional Review Board (IRB) requirements.

### RNA Extraction from human retinal tissues

Total RNA was extracted from approximately 10 mg of human retinal tissue using TRIzol reagent (Invitrogen, Thermo Fisher Scientific, USA), followed by purification with the PureLink RNA Mini Kit (Invitrogen, Thermo Fisher Scientific, USA), including on-column DNase digestion. Retinal tissue (∼10 mg) was homogenized in 800 µL of TRIzol for 30 seconds using a homogenizer (Scilogex D-160, USA), followed by phase separation with 160 µL of chloroform. RNA was subsequently purified using spin columns and eluted in 50 µL of RNase-free water. RNA concentration and purity were assessed using a NanoDrop spectrophotometer (Thermo Fisher Scientific, USA), and samples with A260/A280 ratios between 1.7 and 2.1 were selected for downstream experiments. RNA integrity was evaluated using the RNA ScreenTape assay on a 2200 TapeStation system (Agilent Technologies, USA) and RNA RIMs were shown in Table S2.

### Human iPSC Culture and RPE Differentiation

Human-induced pluripotent stem cells hiPSC line EP1 were cultured and differentiated into RPE monolayers following established protocols. ^20^ Briefly, hiPSCs were maintained on growth factor–reduced Matrigel (BD Biosciences) in mTeSR1 medium (Stem Cell Technologies) under 10% CO₂ and 5% O₂, with expansion promoted by 5 μM blebbistatin (MilliporeSigma). For differentiation, cells were seeded at 35,000 cells/cm² in mTeSR1 until confluency, then switched to differentiation medium (DMEM/F12 supplemented with 15% knockout serum, 2 mM glutamine, 1% nonessential amino acids, 0.1 mM mercaptoethanol, 1% antibiotic–antimycotic, and 10 mM nicotinamide, all are from Thermo Fisher Scientific, USA) for ∼50 days. Differentiating RPE cells were dissociated with 0.25% collagenase IV (Thermo Fisher Scientific, USA) and AccuMAX (MilliporeSigma, USA) to generate a single-cell suspension, then re-plated on Matrigel-coated plates. Cells were matured for 2–3 months in RPE medium (70% DMEM, 30% Ham’s F12, 2% B-27 supplement, and 1% antibiotic–antimycotic, all were from Thermo Fisher Scientific, USA).

### Cigarette Smoke Extract (CSE) and paraquat treatments for iPSC-derived RPE monolayers

Human iPSC-derived RPE monolayers were differentiated using our established methods and cultured for eight weeks to achieve mature RPE cultures. ^21^ The mature monolayers were then treated for 48 hours with Cigarette Smoke Extract (CSE) (Murty Pharmaceuticals, USA) and paraquat (MilliporeSigma, USA) at three lethal dose (LD) concentrations: LD25, LD50, and LD75 (doses for CSE: 125, 250, and 500 μg/mL, respectively; doses for PQT: 100, 300, and 600 μM, respectively). The CSE was prepared in DMSO, with the final solution containing 40mg/mL condensate. Following treatment, total RNA was extracted from the RPE monolayers using RNeasy Mini Kit (QIAGEN, USA). RNA purity and concentration were assessed using NanoDrop™ spectrophotometry (Thermo Fisher Scientific, USA), and only samples with A260/A280 ratios between 1.8 and 2.1 were used for downstream qPCR analysis.

### Reverse transcription and Quantitative PCR (qPCR)

Total RNA (500 ng) isolated from human retinal tissues or induced pluripotent stem cell-derived retinal pigment epithelial (iPSC-RPE) cells was reverse-transcribed into cDNA using either the High-Capacity cDNA Reverse Transcription Kit (Applied Biosystems, USA) or the iScript™ cDNA Synthesis Kit (Bio-Rad, USA), according to the manufacturers’ instructions. Reverse transcription was performed in a total reaction volume of 20 µL, and the resulting cDNA was diluted to a final volume of 60 µL. Quantitative PCR (qPCR) was performed using the CFX384 Real-Time PCR Detection System (Bio-Rad, USA) with iTaq Universal SYBR Green Supermix (Bio-Rad, USA). Each 5 µL reaction contained 0.5 µL of diluted cDNA (equivalent to approximately 6.25 ng of input total RNA), 200 nM of each primer, and 1× SYBR Green Supermix. The thermal cycling conditions consisted of an initial denaturation at 95°C for 30 s, followed by 39 cycles of denaturation at 95°C for 5 s and annealing/extension at 60°C for 20 s. All experiments were performed using three biological replicates.

Relative gene expressions were calculated using the geometric mean of two housekeeping genes, GAPDH and 18S (use a 100-fold dilution of 18S cDNA for qPCR analysis), for normalization. Primer sequences are detailed in Supplementary Table S1 and Figure 2B-C.

### RNA-sequencing (RNA-seq) and data analysis

Published RNA-seq datasets were re-analysed for expression of genes within the *PLEKHA1–ARMS2–HTRA1* locus. ^22^ Reads were mapped to the region of interest and quantified using standard pipelines.

Raw RNA-seq data associated with the study by Wang J et al.^22^ was obtained from the Gene Expression Omnibus under the accession reported in the original publication. FASTQ files were subjected to quality assessment using FastQC, followed by adapter and low-quality base trimming with Trim Galore. High-quality reads were aligned to the human reference genome (GRCh38) using STAR (v2.7 or later) with default parameters, and uniquely mapped reads were retained for downstream analysis. Gene-level read counts were generated using featureCounts based on GENCODE comprehensive gene annotation (v3x). Differential gene expression analysis between age-related macular degeneration (AMD) and control retinal or RPE/choroid samples was performed in R (v4.x) using the DESeq2 package. FPKM values were normalized from the raw fragment counts. The counts were derived from sequencing fragments. When a fragment is large, it appears as a line in the figure. Otherwise, it is displayed as scattered dots. Statistical significance was determined using the Wald test with Benjamini–Hochberg correction (adjusted p < 0.05). For selected genes (*ARMS2* and H*TRA1*) and loci of interest, transcript-level quantification was further performed using Salmon (v1.x), and results were visualized using ggplot2.

### Sanger Sequencing

DNA sequencing was performed at the Johns Hopkins Genetics Resources Core Facility using the Applied Biosystems 3730xl DNA Analyzer (Thermo Fisher Scientific, USA).

### Analysis of published retinal scRNA-seq datasets

Publicly available single-cell RNA sequencing (scRNA-seq) datasets of human retina were obtained from the Gene Expression Omnibus (GEO; accession numbers GSE155288, GSE188280, if using excel table HTRA1_AS_counts_v4.xlsx, GSE210543, GSE278749, or GSE203499, depending on what’s included. Raw count matrices and associated metadata were analyzed using the Seurat package (v5.5.1) in R. Cells with gene counts < 190 were excluded. Gene expression values were normalized using LogNormalize, followed by principal component analysis (PCA) and Uniform Manifold Approximation and Projection (UMAP) for dimensionality reduction and visualization. Cell clusters were annotated based on the original publication and established retinal cell-type marker genes. Expression of genes of interest was visualized using ViolinPlot functions in Seurat.

### Software and Statistical Analysis

Data were analysed using GraphPad Prism (GraphPad Software10.3, USA) and Microsoft Excel. Statistical significance was assessed with unpaired two-tailed Student’s t-tests unless otherwise indicated. Data are presented as mean ± SEM (n = biological replicates). ns = not significant; * p < 0.05; ** p < 0.01; *** p < 0.001; **** p < 0.0001. A p-value < 0.05 was considered statistically significant. AlphaFold3 was used for predicting the three-dimensional structures of the putative peptide coding regions.

## Results

### RNA-seq identifies aligned-read peaks overlapping *HTRA1-AS1*

Reanalysis of RNA-seq datasets from macular retina, peripheral retina, and RPE/choroid tissues across the *ARMS2–HTRA1* locus revealed two prominent read-alignment peaks upstream and one downstream of *ARMS2*, consistently detected in both retinal and RPE/choroid samples (Figure 1). *HTRA1* exhibited robust expression across all ocular tissues (100–550 FPKM) with no marked preference between retina and RPE, whereas *ARMS2* expression was minimal and largely confined to the retina (∼5–10 FPKM). The antisense transcript *HTRA1-AS1* (annotated on NCBI as predicted *<u>HTRA1</u>* <u>and *ARMS2* antisense RNA 1</u>) were expressed at modest levels (∼30–50 FPKM), representing only a small fraction of *HTRA1* expression. These antisense peaks correspond to low-abundance noncoding transcription within the 10q26 risk interval, supporting the presence of additional regulatory elements in this region.

**Figure 1.**
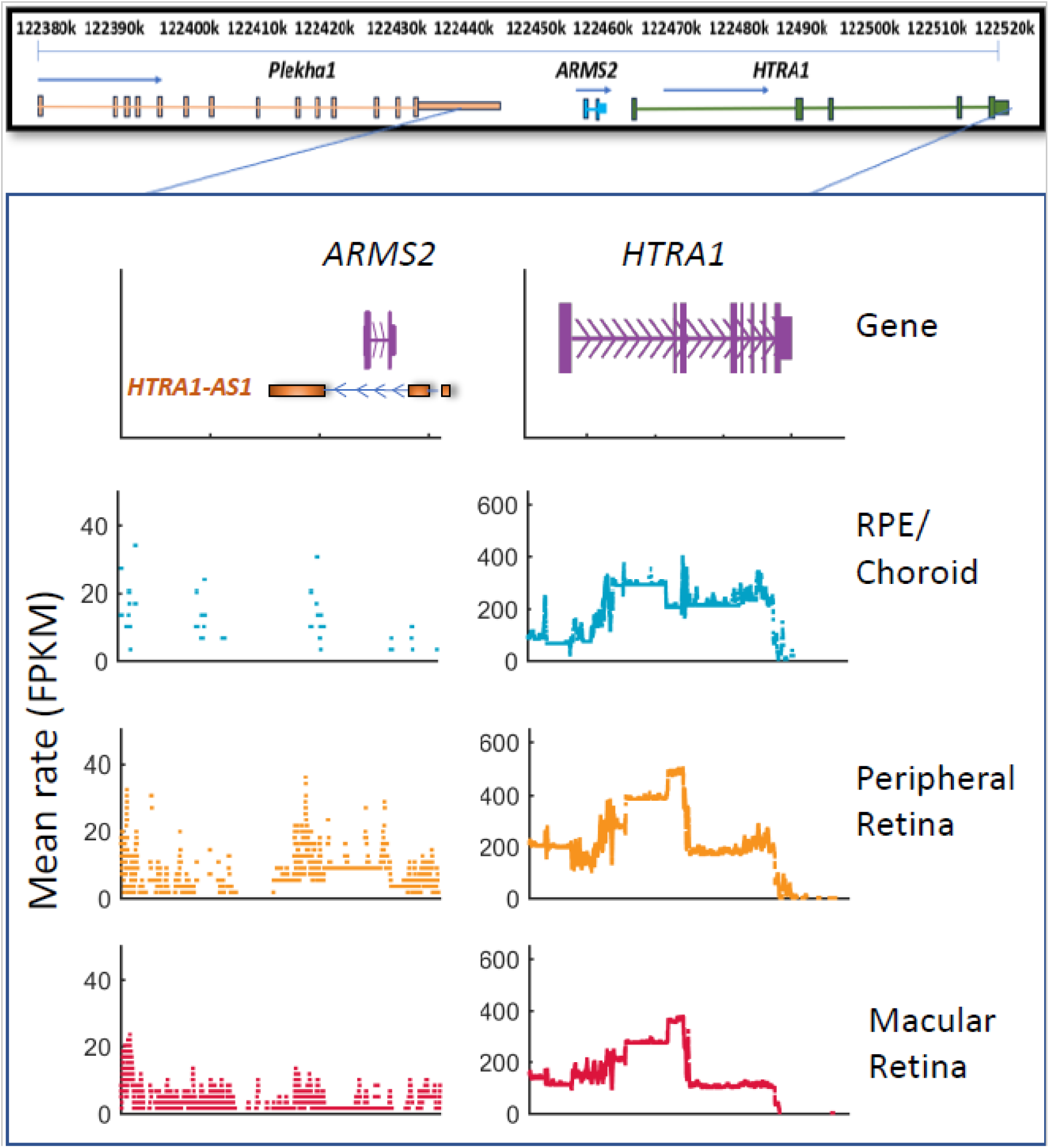
Genomic organization and expression profiles of *HTRA1-AS1* and *HTRA1* in human retinal tissues. Schematic representation of the genomic organization of *HTRA1-AS1, ARMS2*, and *HTRA1* (upper panels) and RNA sequencing (RNA-seq) read coverage across these regions in human RPE/choroid, peripheral retina, and macular retina (lower panels). The left panels show the genomic location and expression profile of *HTRA1-AS1*, an antisense long noncoding RNA overlapping the *ARMS2* locus, whereas the right panels show the corresponding genomic organization and expression profile of *HTRA1*. RNA-seq read coverage is presented as mean fragments per kilobase of transcript per million mapped reads (FPKM). Blue, orange, and red indicate RPE/choroid, peripheral retina, and macular retina, respectively. *HTRA1-AS1* exhibited relatively low expression across the examined tissues, whereas HTRA1 showed substantially higher expression levels. These findings demonstrate distinct expression profiles of *HTRA1-AS1* and *HTRA1* across human retinal tissues.

### Validation of lncRNA *HTRA1-AS1* and transcript variants

Database searches identified multiple long non-coding RNAs (lncRNAs) overlapping *ARMS2*. Ensembl listed four candidates (*ENST00000647969.1, ENST00000650300.1, ENST00000811415.1,* and *ENST00000811416.1*), and NCBI annotated *HTRA1-AS1* (*LOC105378525*) with four predicted transcript variants (XR_946382–946385). All were transcribed antisense to *ARMS2* (Figure 2A).

We developed an optimized strategy for designing PCR primers specific to five RNA variants, including *HTRA1-AS1*, *ENST00000647969.1, ENST00000650300.1, ENST00000811415.1,* and *ENST00000811416.1*. Under refined experimental conditions, this approach yields a single, specific band for each variant, the identities of which were validated via Sanger sequencing (Figure 2B-D). A comprehensive description of this methodology is available as a preprint (Ahn et al. Zack DJ, Zhang PW, BioRxiv, 2026). ^23^While all five variants can be amplified using high PCR cycle numbers, a primer pair originally intended to co-amplify *ENST00000647969.1*, *ENST00000811415.1, ENST00000811416.1*, and *HTRA1-AS1* produced only three distinct bands (Figure 2E). Gel extraction and Sanger sequencing identified these as *ENST00000811416.1(*305 bp) and *HTRA1-AS1(*481 bp). The intermediate band (∼400 bp) was also observed; however, denaturation and re-annealing experiments confirmed that it represents a heteroduplex formed between the two primary products (Figure 3 of the referenced preprint).^23^

Subsequent analyses indicated that *ENST00000811416.1* and HTRA1-AS1 are the dominant variants in this system; however, their relative abundances could not be definitively resolved in three non-AMD control and three AMD retinal samples. (Figure 2F). In contrast, *ENST00000650300.1* exhibited minimal expression (qPCR Ct ∼35). Consequently, a qPCR primer pair that co-amplifies *HTRA1-AS1 and ENST00000811416.1* was employed to assess their combined expression levels, representing the transcribed RNAs of the *HTRA1-AS1* gene.

**Figure 2.**
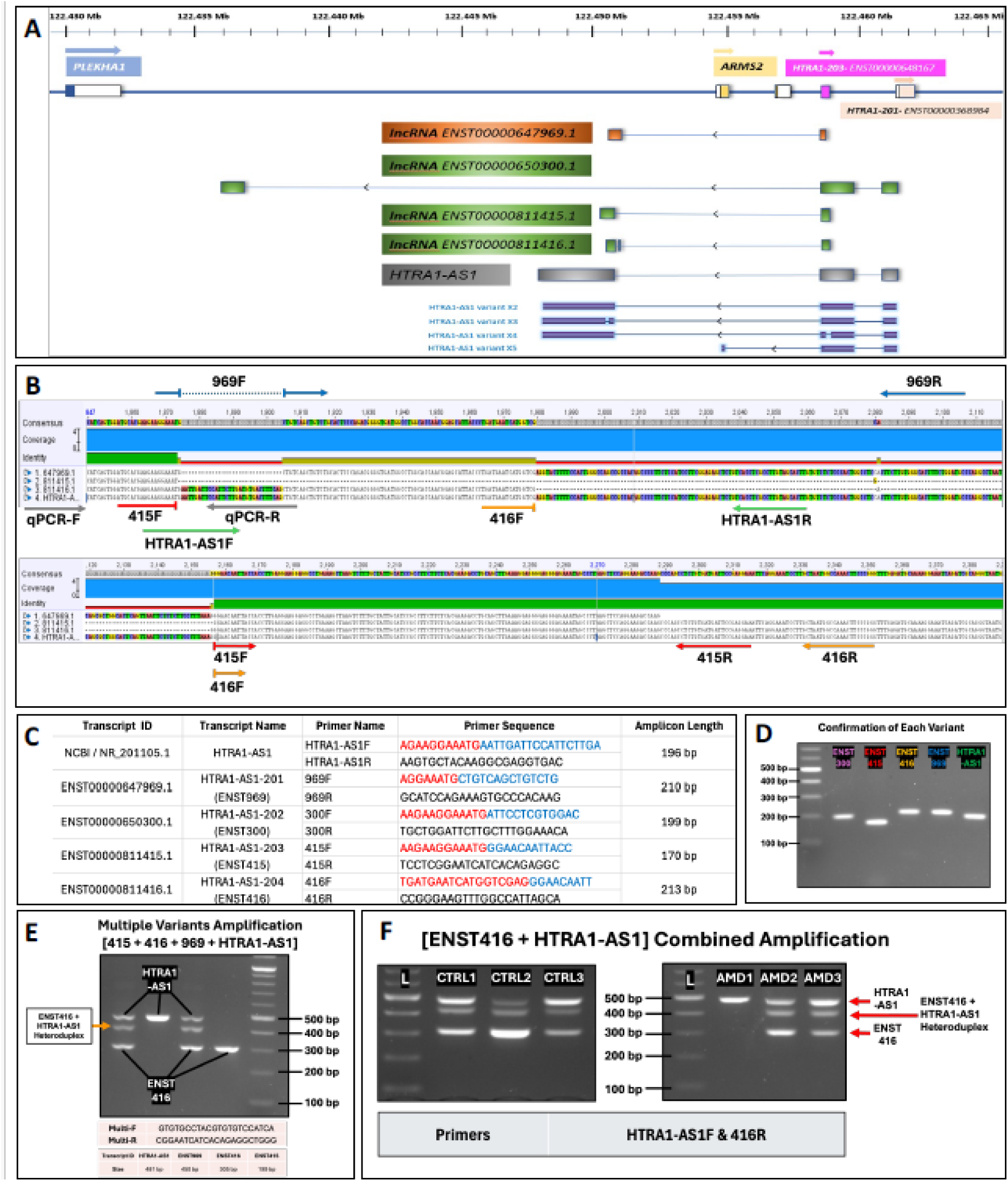
Schematic illustrating the overlap of the association peak with the *ARMS2* locus and overlapping lncRNA variants. (**A**) Genomic organization of lncRNA candidates within the *PLEKHA1–ARMS2–HTRA1* region on chromosome 10q26, showing annotated protein-coding genes (blue, yellow, and coral) and multiple lncRNAs (shaded in green, orange, and gray). Coordinates are indicated in megabases (Mb). Exons are represented by boxes and introns by connecting lines; arrows indicate transcriptional direction. (**B**) Alignment of four lncRNAs (*ENST00000647969.1, ENST00000811415.1, ENST00000811416.1 and HTRA1-AS1*), with primer positions indicated for variant-specific amplification. (**C**) Variant identification and primer sequences for exon–exon junction RT-PCR. Primer sequences are provided, with forward primers color-coded to indicate their exon-binding regions. (**D**) Variant-specific amplification using exon–exon junction RT-PCR. (**E)** Multiplex amplification of four variants; expected product sizes are indicated. (F) Co-amplification of *ENST00000811416.1* and *HTRA1-AS1* in non-AMD control (CTRL) and AMD retinal samples

### *HTRA1-AS1* is downregulated in human AMD retinas

To assess the relationship between lncRNA *HTRA1-AS1* expression and age-related macular degeneration (AMD), we collected human AMD and non-AMD control donor eyes from the National Disease Research Interchange (NDRI) over a period of more than five years. Most donors were White, and the two groups were age- and sex-matched. Sex distribution was also comparable (control: 19 males, 24 females; AMD: 22 males, 21 females). Representative images of control and AMD eyes are shown in Figure 3A-B.

*HTRA1-AS1* was significantly downregulated in AMD eyes (p = 0.007; Figure 3C). In contrast, *HTRA1* mRNA levels did not differ significantly between AMD and controls (p = 0.121; Figure 3D). *ARMS2* expression was extremely low (Ct >34.8) in our samples and not reliably quantifiable.

**Figure 3.**
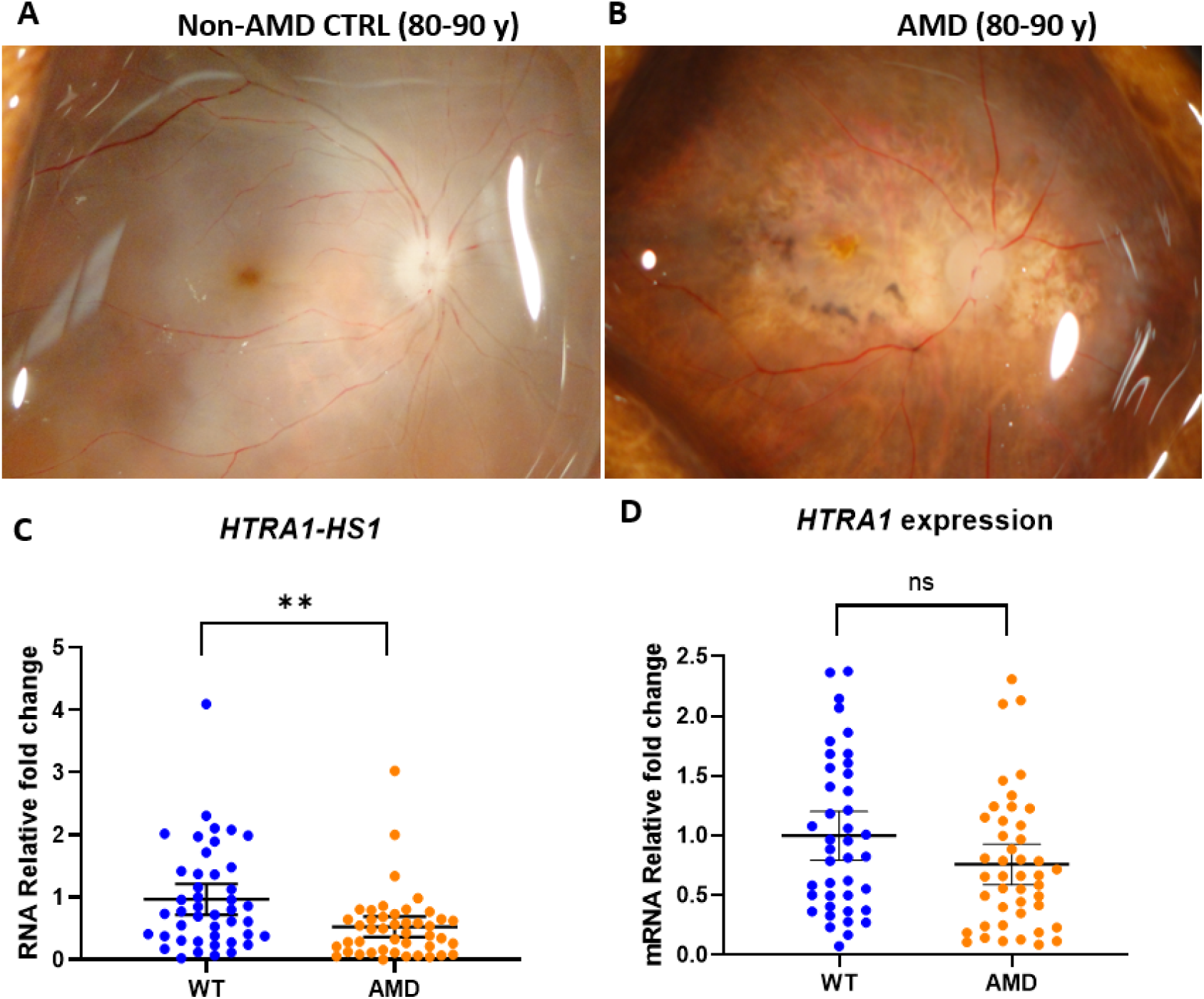
RT-qPCR analysis of RNA expression in control and AMD donor retinas. (**A, B**) Representative macular images from non-AMD control and AMD eyes. (**C, D**) *HTRA1-AS1* and *HTRA1* RNA expression. Data was presented as the mean ± 95% confidence interval (CI)

### Cell-type expression profiles of *HTRA1-AS1* and *HTRA1*

Single-cell RNA sequencing (scRNA-seq) analysis of human retinal tissues revealed distinct cell-type-specific expression patterns of *HTRA1-AS1* and *HTRA1* in both macular and peripheral retinal regions (Figure 4). *HTRA1-AS1* expression was predominantly detected in bipolar cells, rod photoreceptors, and Müller glial cells, with relatively low expression in amacrine cells, cone photoreceptors, microglia, pericytes, astrocytes, horizontal cells, ganglion cells, and endothelial cells (Figure 4A). In contrast, *HTRA1* exhibited a distinct expression pattern, with the highest expression levels observed in horizontal cells and detectable expression in several other retinal cell populations, including rods, Müller glia, and astrocytes (Figure 4C). Further analysis of regional expression revealed that both *HTRA1-AS1* and *HTRA1* were detectable in the macular and peripheral neural retina and choroid/RPE (Figures 4B and 4D). HTRA1-AS1 expression was relatively low and restricted to subsets of cells across these regions, whereas HTRA1 exhibited a broader expression distribution. Together, these findings demonstrate distinct cell-type-specific expression profiles of *HTRA1-AS1* and *HTRA1* in the human retina, despite their overlapping regional expression patterns.

**Figure 4.**
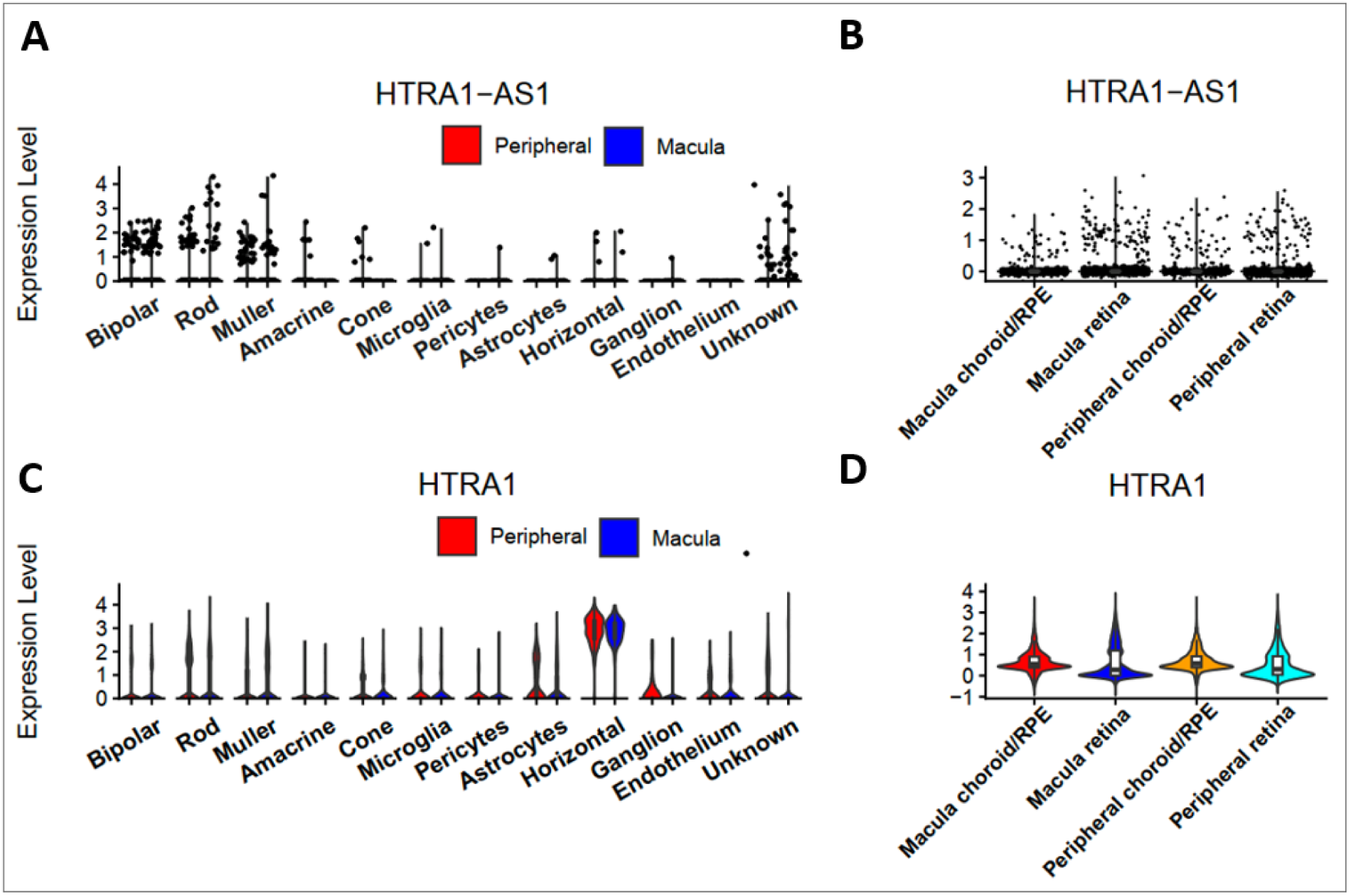
Cell-type-specific and regional expression patterns of *HTRA1-AS1* and *HTRA1* in the human retina. Single-cell RNA sequencing (scRNA-seq) analysis showing the expression patterns of *HTRA1-AS1* and *HTRA1* across different retinal cell types and anatomical regions. (A) *HTRA1-AS1* expression across retinal cell populations, including bipolar cells, rods, Müller glia, amacrine cells, cones, microglia, pericytes, astrocytes, horizontal cells, ganglion cells, endothelial cells, and unclassified cells (Unknown). (B) *HTRA1-AS1* expression in macular choroid/RPE, macular retina, peripheral choroid/RPE, and peripheral retina. (C) *HTRA1* expression across the same retinal cell populations, with the highest expression observed in horizontal cells. (D) *HTRA1* expression in the four anatomical regions shown in panel B. Red and blue indicate peripheral and macular samples, respectively, in panels A and C. Each dot in panels A and B represents an individual cell, whereas violin plots in panels C and D illustrate the distribution of expression levels across cells. *HTRA1-AS1* was predominantly detected in bipolar cells, rods, and Müller glia, whereas *HTRA1* exhibited a distinct expression pattern characterized by high expression in horizontal cells

### Cigarette smoke extract (CSE), but not paraquat (PQT), selectively induces dose-dependent upregulation of *HTRA1-AS1* and *ARMS2* mRNA

Because cigarette smoking is a major risk factor for AMD, we investigated the transcriptional response of *HTRA1-AS1* and neighbouring genes to CSE in iPSC-derived RPE cells. CSE treatment elicited a robust, dose-dependent induction of *HTRA1-AS1*, and *ARMS2* transcripts, with the highest increases observed at the LD75 dose (8.0-fold, and 75.3-fold, respectively; Figure 5A, 5C). *HTRA1* mRNA expression also rose at higher CSE concentrations, though to a lesser extent, reaching a maximum 2.8-fold increase (Figure 5B).

To test if this response was specific to CSE or a general reaction to oxidative stress, we treated cells with paraquat (PQT), a classic oxidative stress inducer. Paraquat is a fast-acting, non-selective herbicide that induces toxicity through redox cycling, generating reactive oxygen species (ROS) and causing oxidative damage to cellular components. It is widely used in experimental models to study oxidative stress and neurodegeneration, including features of Parkinson’s disease. In contrast, PQT treatment across comparable sublethal concentrations (LD25–LD75) did not significantly alter the expression of any of the tested transcripts (Figure 5D–G). These findings demonstrate that CSE selectively drives the dose-dependent induction of *HTRA1-AS1* and *ARMS2* in RPE cells, suggesting a smoking-specific regulatory effect on this AMD-associated locus.

**Figure 5.**
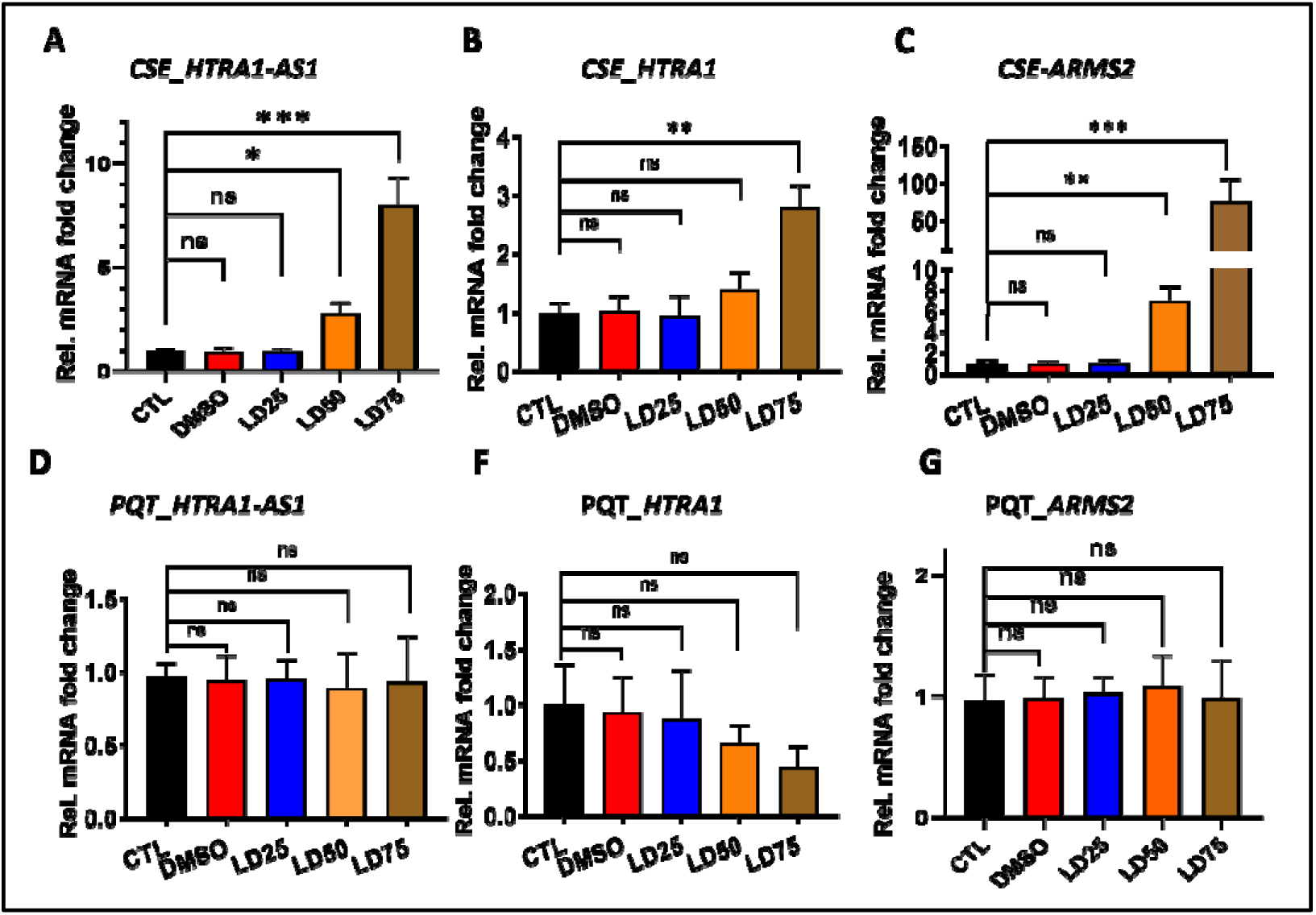
Quantitative RT-PCR analysis of the expression of *HTRA1-AS1, HTRA1* and *ARMS2* mRNA in cells following treatment with increasing concentrations of CSE (Panels A–C) or PQT (Panels D–G). Treatment groups included untreated control (CTL), vehicle control (DMSO), and three lethal dose (LD) levels: LD25, LD50, and LD75. mRNA levels were normalized to GAPDH and 18S. (A–D). Data shown corresponds to technical replicates from one biological replicate, with findings independently validated in a second biological replicate. The Ct value for ARMS2 expression in retina was >34.8. In untreated iPSC-RPE cells, the Ct values for *ARMS2, HTRA1-AS1 and HTRA1* were 31.7, 28.4 and 23.7, respectively

## Discussion

A major challenge in AMD research is reconciling the strong genetic associations at the *ARMS2/HTRA1* locus with limited molecular evidence supporting a causative mechanism. Previous investigations have largely focused on the protein-coding genes *ARMS2* and *HTRA1*.²⁴ In the present study, we broadened this scope to include noncoding transcripts within the chromosome 10q26 risk locus, which has been consistently implicated in AMD by genome-wide association studies (GWAS). Importantly, RNA read densities across this region often exceeded those mapped to *ARMS2* exons, suggesting that additional regulatory elements beyond *ARMS2* and *HTRA1* may contribute to disease risk. Traditional approaches to identifying disease-associated genes include candidate gene analyses, linkage studies combined with whole-exome or whole-genome sequencing in familial disorders, and GWAS followed by functional investigations in complex diseases. By integrating retinal RNA-seq data with NCBI and Ensembl annotations, we identified and characterized the antisense long noncoding RNA (lncRNA) *HTRA1-AS1* and its transcript variants, which overlap the *ARMS2* gene.

Although several lncRNAs have been implicated in AMD,²⁵˒²⁶ none of the previously described lncRNAs were initially identified within the *ARMS2/HTRA1* risk locus. A recently published study reported that the same lncRNA (BX842242.1/ENSG00000285955) is associated with reticular pseudodrusen (RPD), with elevated *HTRA1-AS1* expression correlating with reduced *HTRA1* levels.²⁷ In contrast, our data demonstrate that *HTRA1-AS1* is significantly downregulated in AMD retinas, whereas *HTRA1* mRNA expression does not differ significantly between cases and controls. These apparently divergent findings may reflect differences in disease phenotypes, tissue composition, retinal regions examined, or analytical approaches. They also suggest that the relationship between *HTRA1-AS1* and *HTRA1* expression may be context-dependent rather than governed by a simple inverse regulatory mechanism.

Since the original Yang Z et al reports identifying the *ARMS2/HTRA1* region as an AMD susceptibility locus in 2006, ²⁸ attempts to directly link *ARMS2* or *HTRA1* expression with AMD have yielded inconsistent results, often due to limited sample sizes.¹⁹˒²⁹ By analyzing 43 AMD and 44 control retinas, our study provides evidence of lncRNA dysregulation at this locus in a relatively large cohort of human retinal donors. Specifically, *HTRA1-AS1* was reduced in AMD retinas but was robustly induced by cigarette smoke extract (CSE) in iPSC-derived RPE cells. Interestingly, this effect was not observed with paraquat, a general oxidative stressor, suggesting that *ARMS2* and *HTRA1-AS1* do not respond uniformly to different oxidative stress stimuli. Instead, their selective upregulation by CSE may reflect the involvement of additional molecular pathways activated by components of cigarette smoke. The modest but significant increase in *HTRA1* expression further supports a broader transcriptional response involving multiple genes at this locus. Together, these findings suggest that smoking may influence AMD-associated transcriptional responses at the *ARMS2/HTRA1* locus, potentially contributing to interactions between environmental exposures and genetic susceptibility.

Single-cell RNA-seq analysis further revealed distinct cell-type-specific expression patterns of *HTRA1-AS1* and *HTRA1* in the human retina. *HTRA1-AS1* was predominantly detected in bipolar cells, rod photoreceptors, and Müller glia, whereas *HTRA1* expression was highest in horizontal cells. Both transcripts were detectable in macular and peripheral neural retina and choroid/RPE tissues, although *HTRA1-AS1* exhibited a more restricted cellular expression pattern. These observations suggest that the two transcripts may have distinct regulatory or functional roles despite their genomic proximity. The expression of *HTRA1-AS1* in rod photoreceptors and Müller glia is particularly relevant because these cell populations contribute to retinal function and homeostasis. However, the potential relationship between their expression patterns and AMD-associated cellular dysfunction remains to be determined.

We note that the apparent enrichment of transcripts in specific cell types should be interpreted with caution. Single-cell RNA-seq data are inherently subject to sampling bias, differences in cell capture efficiency, and variability in sequencing depth, which can influence the detection and apparent abundance of transcripts across cell populations. In addition, dropout events and uneven representation of cell types may further skew expression patterns. Therefore, while our data suggests cell-type-specific expression patterns, these observations require further validation using complementary approaches, such as RNA in situ hybridization, and larger, well-balanced datasets.

We also examined whether AMD-associated genetic variants at the chromosome 10q26 locus were associated with *HTRA1-AS1* expression. No significant genotype-dependent differences were detected in our cohort. This finding contrasts with the study by Farashi et al. ^27^, which reported an expression quantitative trait locus (eQTL) effect on *HTRA1-AS1* expression in independent donor retinal cohorts. This discrepancy may reflect differences in cohort size, tissue composition, anatomical regions examined, transcript quantification methods, or statistical power. Importantly, the absence of a statistically significant association in our cohort does not exclude a genotype-dependent regulatory effect, particularly if such effects are modest or restricted to specific retinal cell populations. These findings suggest that the relationship between AMD-associated genetic variants and *HTRA1-AS1* expression may be more complex than previously recognized.

The precise functional role of *HTRA1-AS1* remains to be elucidated. Although antisense lncRNAs frequently regulate the transcription or stability of neighboring genes,²⁴˒²⁵ our data do not support a simple inverse relationship between *HTRA1-AS1* and *ARMS2* expression under the experimental conditions examined. Increasing evidence also indicates that some lncRNAs can produce bioactive micropeptides.³⁰ For example, myoregulin (MLN), originally annotated as the long noncoding RNA LINC00948, was later identified as a 46-amino-acid transmembrane micropeptide. MLN directly interacts with SERCA to inhibit Ca²⁺ uptake into the sarcoplasmic reticulum, thereby regulating calcium homeostasis and muscle contractile function.³¹ Notably, the AMD-associated SNP rs61871745 (G/A; A as the risk allele) resides within putative open reading frames (ORFs) of both *HTRA1-AS1* and *ENST00000647969.1*. The risk allele (A) is predicted to result in a codon substitution from ACC to ATC (Thr→Ile) within a putative ORF (Figures 6A–B), although *ENST00000647969.1* exhibits minimal expression in the present study. Structural modeling suggests that this amino acid substitution could influence the properties of the predicted peptide (Figure 6C). Although sequence analysis identified putative open reading frames (ORFs) within *HTRA1-AS1* transcripts that could potentially encode micropeptides, our examination of publicly available datasets and repositories did not reveal experimental evidence supporting their translation under physiological conditions. Thus, the proposed micropeptide-coding potential of *HTRA1-AS1* remains speculative. Further studies, including ribosome profiling, targeted proteomics, and functional assays, are needed to establish whether these transcripts are translated and whether their putative micropeptides play functional roles in retinal physiology or AMD pathogenesis. Whether *HTRA1-AS1* functions solely as a regulatory lncRNA, encodes a functional micropeptide, or possesses dual roles remains an important question for future investigation.

In addition to the well-characterized degeneration of photoreceptors and retinal pigment epithelium (RPE), choroidal vascular alterations and degeneration are important components of AMD pathology. The choroid is essential for maintaining outer retinal homeostasis by supplying oxygen and nutrients to photoreceptors and facilitating metabolic waste removal. Degeneration of the choriocapillaris and associated vascular structures can result in impaired perfusion, hypoxia, and reduced support for the overlying RPE and photoreceptors, thereby exacerbating retinal dysfunction. These changes are often accompanied by alterations in extracellular matrix composition and increased inflammatory signaling within the choroid, further contributing to disease progression.³²˒³³˒³⁴ Given our detection of *HTRA1-AS1* in both macular and peripheral choroid/RPE tissues, future studies should investigate its expression and potential function in these compartments, particularly in relation to choroidal vascular alterations and AMD progression.

Risk alleles at the *ARMS2/HTRA1* locus have been consistently associated not only with susceptibility to age-related macular degeneration but also with the progression of geographic atrophy (GA). Several studies have reported that individuals carrying these risk variants exhibit faster enlargement of atrophic lesions, suggesting a role in disease severity and progression.³⁵˒³⁶ Although the precise mechanisms remain unclear, these variants may influence extracellular matrix remodeling, inflammatory responses, and choroidal integrity, thereby exacerbating degeneration of the RPE and overlying photoreceptors. Whether *HTRA1-AS1* contributes to these processes remains unknown. Nevertheless, its localization within this genetically important region and its altered expression in AMD retinas warrant further investigation into its potential involvement in disease progression.

An additional limitation of this study is the lack of detailed clinical staging information for the AMD donor eyes. Information regarding disease stage (early, intermediate, or advanced AMD) would provide important context, particularly given the known associations of *ARMS2/HTRA1* with disease progression and geographic atrophy. Unfortunately, detailed clinical staging information was not provided by the National Disease Research Interchange (NDRI). However, based on our preliminary assessment of ocular morphology, we estimate that most AMD donor samples correspond to early- to intermediate-stage dry AMD. Because this classification was not confirmed by comprehensive clinical records, we could not reliably assess the relationship between *HTRA1-AS1* expression and AMD stage or specific disease phenotypes.

In summary, our study identifies and characterizes *HTRA1-AS1* and its transcript variants as antisense lncRNAs overlapping the ARMS2 locus and demonstrates their altered expression in AMD retinas. The distinct cell-type-specific expression patterns, disease-associated downregulation, and responsiveness to CSE-induced stress suggest that *HTRA1-AS1* may participate in molecular pathways relevant to AMD pathogenesis. Although its regulatory functions and potential micropeptide-coding capacity remain to be experimentally established, these findings expand our understanding of the transcriptional complexity of the chromosome 10q26 risk locus. Pinpointing the causal variants and clarifying the potential regulatory and/or coding roles of *HTRA1-AS1* will be important for elucidating the molecular mechanisms underlying the association between the *ARMS2/HTRA1* locus and AMD susceptibility and may ultimately identify new targets for therapeutic investigation.

**Figure 6.**
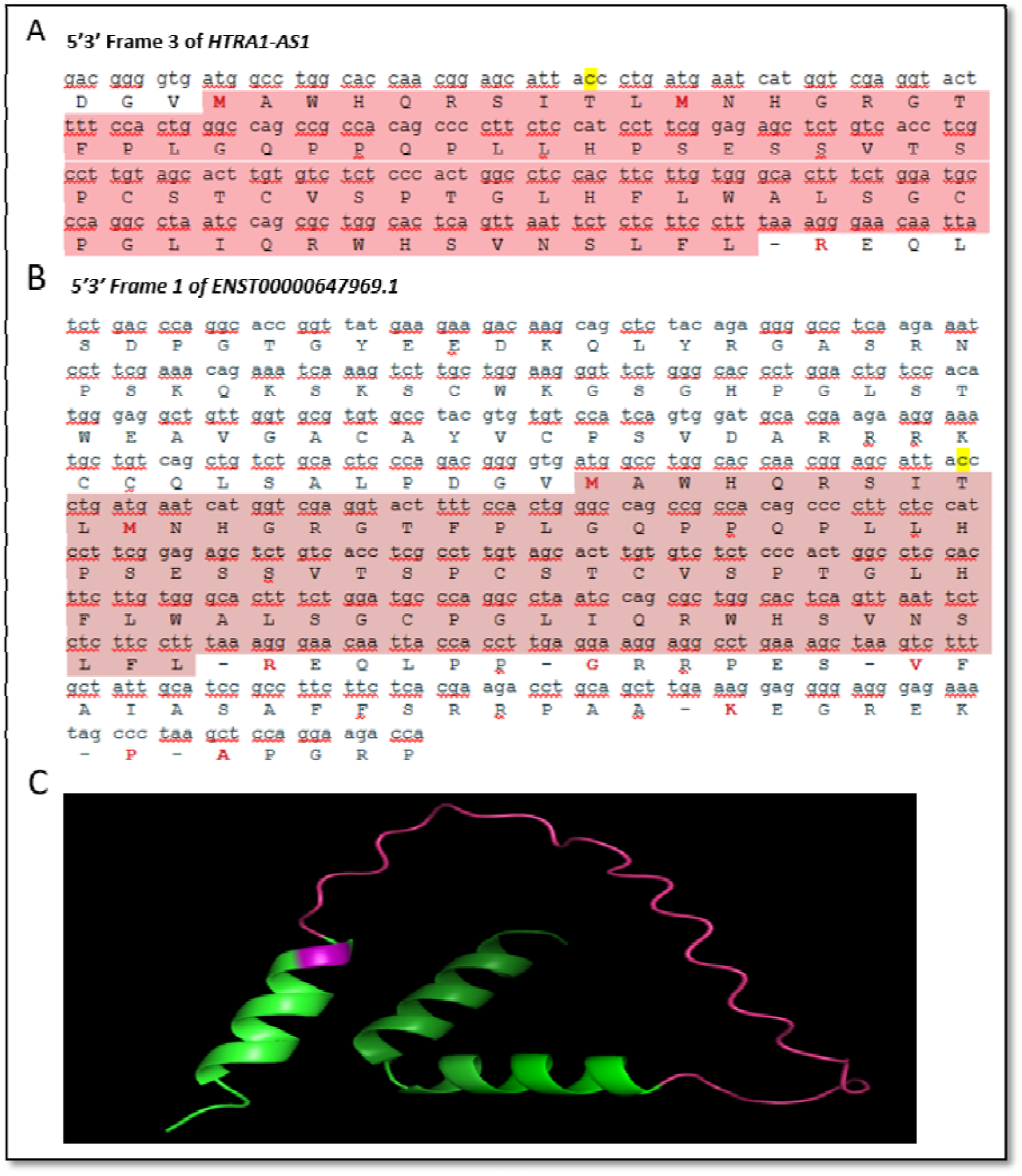
Predicted coding sequences and protein structure for *HTRA1-AS1* and *ENST00000647969.1*. **A and B**, Nucleotide and corresponding amino acid sequences of the putative open reading frames (ORFs) identified in the 5′–3′ Frame 3 of *HTRA1-AS1* (**A)** and the 5′–3′ Frame 1 of *ENST00000647969.1* (**B**). Coding regions are shaded in red (start codon to stop codon) and the rs61871745 is highlighted in yellow. (**C**) Predicted three-dimensional structures of the peptides encoded by the putative ORFs in *HTRA1-AS1* and *ENST00000647969.1*, generated using AlphaFold. The structures display alpha-helices and loop regions, with the amino acid change caused by SNP rs61871745 indicated in pink.

## Data Availability

All data produced in the present work are contained in the manuscript

## Contributors

PWZ conceived the study, designed all experiments, and wrote the primary manuscript draft. SL performed the bioinformatic screening for trans-acting element binding. ZHW, WL, JW, SS, JA,SL, LF, JW, CAB, and JQ contributed substantially to experimental execution, data collection, and analysis. PWZ, SLM, and DJZ supervised the overall process of data acquisition, analysis, interpretation, and manuscript preparation.

## Data sharing

The authors declare that all data generated or analysed during this study are included in this published article.

## Acknowledgement

The authors thank Dr. James T. Handa for his insightful suggestions and discussions, and Dr. Jikui Shen for his valuable assistance in predicting the three-dimensional structures of the putative peptide coding regions using AlphaFold. This study was supported by NEI P30 EY001765 (Wilmer Core Grant, Microscopy Module), NEI R01EY020406 and NEI X01HG006605.

