## supplementary file for "*HTRA1-AS1*, an *ARMS2*-region long non-coding RNA, is downregulated in retinas of age-related macular degeneration patients"

**Supplementary data**

1. **Table S1 Primer sequences used for quantitative PCR (qPCR)**

| **qPCR Primers** | **Primer sequences** |
| --- | --- |
| hGAPDH-F | CTGACTTCAACAGCGACACC |
| hGAPDH-R | TAGCCAAATTCGTTGTCATACC |
| hHTRA1-AS1-qPCR-F | TCACCCCACCTGCTCTGACC |
| hHTRA1-AS1-qPCR-R | ACAGCTGAAAGTCACATCAAGAATGGA |
| hHTRA1-F | AAGCCAAAGAGCTGAAGGACCG |
| hHTRA1-R | CTTCATTACCCCTGCGGACCAC |
| hARMS2-F | TGACAAGCAGAGGAGCAAACTGT |
| hARMS2-R | TGTTTTGGCAGAATCACCTTGCTG |
| h18SrRNA-F | CAGCCACCCGAGATTGAGCA |
| h18SrRNA-R | TAGTAGCGACGGGCGGTGTG |

Note: Forward (F) and reverse (R) primers were designed for the detection of human *GAPDH* (housekeeping control), *ENST00000647969.1* (lncRNA isoform), *HTRA1-AS1* (lncRNA), *HTRA1*, and ARMS2 transcripts. All sequences are listed in the 5′ to 3′ direction.

2. **The position of rs61871745 in long non-coding RNAs**

**SNP Rs61871745 (original G/A, antisense** C/T) position is highlighted in yellow.

**LncRNA *HTRA1-AS1* (LOC105378525 = NR_201105.1)**

**5'3' Frame 1**

```
ggg cgc gcc cgg gca cgg cgg tgc tgg cgg ggc gcg ggc ggg gca gcg cac agg ccg gcc
 G   R   A   R   A   R   R   C   W   R   G   A   G   G   A   A   H   R   P   A
ctc tcg cgg gac tca gtt tcc ccg tct gcg cgc ccc aca tat tcg ctt tcc gca ggt tct
 L   S   R   D   S   V   S   P   S   A   R   P   T   Y   S   L   S   A   G   S
ctc gct gag att cgc gtc ctt caa act aat gga act ttg caa ggg ggc cct gcg gcg gct
 L   A   E   I   R   V   L   Q   T   N   G   T   L   Q   G   G   P   A   A   A
tcg cgt ggg agc gcc ggg gaa agt tcc tgc aaa tcg ccg gac tgg ggg ccc gcc cgg gaa
 S   R   G   S   A   G   E   S   S   C   K   S   P   D   W   G   P   A   R   E
gct cgg act ggg ccg ggc agg gac tgc agg ggc ctc tgc gca gac ccg ggg cgt tgc ggc
 A   R   T   G   P   G   R   D   C   R   G   L   C   A   D   P   G   R   C   G
acc gcg gac ccc ggt cac gcg ctg gtt ctg ccc ccg gta ccc cgc acg cgg gac cct gac
 T   A   D   P   G   H   A   L   V   L   P   P   V   P   R   T   R   D   P   D
cgc ggg gcc tct gcg gcc gga cga agg cag cgt ccg cgg agc tgg cag aaa gcc cgc gcc
 R   G   A   S   A   A   G   R   R   Q   R   P   R   S   W   Q   K   A   R   A
cag acc cac agt gaa gtg ata aaa gca gag agc aga aaa act gag ggc cag aaa ggg tga
 Q   T   H   S   E   V   I   K   A   E   S   R   K   T   E   G   Q   K   G   -
ctc ccc tga gtg tca tgc cac aag ttg eag tgc tgg agc cag gac acc ttc caa acc ggc
 L   P   -   V   S   C   H   K   L   X   C   W   S   Q   D   T   F   Q   T   G
tgt gcc tgc ctg caa gcc cca tcc tcc tcc atc ccc ctg agc agc cag act gca gag gga
 C   A   C   L   Q   A   P   S   S   S   I   P   L   S   S   Q   T   A   E   G
atg agc gga ccg aaa aca gtg tgg ccg caa ctt ccc cac tgc ttt gcg gga ggt gga ggg
 M   S   G   P   K   T   V   W   P   Q   L   P   H   C   F   A   G   G   G   G
agt ttc tag gct ctt tcc ctg ttg ggg aat taa gta gct ggg gat ccc act gag tct cca
 S   F   -   A   L   S   L   L   G   N   -   V   A   G   D   P   T   E   S   P
cca ccc tct cac cca ttt tcc acc cct ccc cca cca cat ata cat tct gct ccc cca gtg
 P   P   S   H   P   F   S   T   P   P   P   P   H   I   H   S   A   P   P   V
cca aat aac ttg cat ttt ctg gaa cac acc tgt ctg ctt acg gtc tct gtg cct ttc ttt
 P   N   N   L   H   F   L   E   H   T   C   L   L   T   V   S   V   P   F   F
gcc cac atc tcc cct ctt ctt gaa aca ccc ctc ctc ccc tcc agc gtg ggc tca tgt gac
 A   H   I   S   P   L   L   E   T   P   L   L   P   S   S   V   G   S   C   D
cta gag aag cac gtg gga gcc ctg tca tct ggg gga ctt ggg ttc aaa tcc cac ttc tgg
 L   E   K   H   V   G   A   L   S   S   G   G   L   G   F   K   S   H   F   W
tcc tta agt ctg tga ctc tgg gcc ttt gtt taa taa ctg gag gag ttt cat tag gaa aat
 S   L   S   L   -   L   W   A   F   V   -   -   L   E   E   F   H   -   E   N
ggc agt gac cac ata tgt ccc tgg agt gac tga tgt gcc acc cac aag ctt cta ttt ggt
 G   S   D   H   I   C   P   W   S   D   -   C   A   T   H   K   L   L   F   G
cag tgc tgg gtc cca tcg tcc cat cta gca tgc tcc tct tcc atc ctc gtg ccc cca ctc
 Q   C   W   V   P   S   S   H   L   A   C   S   S   S   I   L   V   P   P   L
agt gcc att gtc tct gcc tct cac cct ctg tgc tgc tat tcc tgc cct cat cac ctc ctc
 S   A   I   V   S   A   S   H   P   L   C   C   Y   S   C   P   H   H   L   L
cct agg gca gag gcc ccg tct tct tta cca tgc ccc agt gcc agc aca gtg ctt tgg caa
 P   R   A   E   A   P   S   S   L   P   C   P   S   A   S   T   V   L   W   Q
gtt att cag cac tgg ggg agt aga gtg tat gtg tgg tgg ggg agg ggg tgg aga atg ggt
 V   I   Q   H   W   G   S   R   V   Y   V   W   W   G   R   G   W   R   M   G
aag agg gtg atg gag act cag ggg gac ccc cag cta ctt aat acc cct gag gaa gtg tcg
 K   R   V   M   E   T   Q   G   D   P   Q   L   L   N   T   P   E   E   V   S
gcc gaa gtg gac cca ctc ctg tga cag ttc aca gcc tag aag agg cgt ctg taa cca cgg
 A   E   V   D   P   L   L   -   Q   F   T   A   -   K   R   R   L   -   P   R
tga cca gta agt gcc agc cct ggg gcc acg tgg ctg ttc tag aga agg aag aca gca cta
 -   P   V   S   A   S   P   G   A   T   W   L   F   -   R   R   K   T   A   L
ccc tga gag cca tgc tgg tgg ctt cat cag gaa ggg agt cag cag ggg gag gac tgg ggg
 P   -   E   P   C   W   W   L   H   Q   E   G   S   Q   Q   G   E   D   W   G
acc aag cgg tcc cta aga agc aga att tgc aat acc caa ggt ttt aga aag agc aac ctg
 T   K   R   S   L   R   S   R   I   C   N   T   Q   G   F   R   K   S   N   L
gca ggg cca ctg cac aat gag atc aca gac tca cct ccc agc tgt gag tgc agc tca ccc
 A   G   P   L   H   N   E   I   T   D   S   P   P   S   C   E   C   S   S   P
cac ctg ctc tga ccc agg cac cgg tta tga aga aga caa gca gct cta cag agg ggc ctc
 H   L   L   -   P   R   H   R   L   -   R   R   Q   A   A   L   Q   R   G   L
aag aaa tcc ttc gaa aca gaa atc aaa gtc ttg ctg gaa ggg ttc tgg gca ccc tgg act
 K   K   S   F   E   T   E   I   K   V   L   L   E   G   F   W   A   P   W   T
gtc cac atg gga ggc tgt tgg tgc gtg tgc cta cgt gtg tcc atc agt gga tgc acg aag
 V   H   M   G   G   C   W   C   V   C   L   R   V   S   I   S   G   C   T   K
aag gaa atg aat tga ttc cat tct tga tgt gac ttt cag ctg tec agc tgt ctg cac tcc
 K   E   M   N   -   F   H   S   -   C   D   F   Q   L   X   S   C   L   H   S
cag acg ggg tga tgg cct ggc acc aac gga gca tta ccc tga tga atc atg gtc gag gta
 Q   T   G   -   W   P   G   T   N   G   A   L   P   -   -   I   M   V   E   V
ctt ttc cac tgg gcc agc cgc cac agc ccc ttc tcc atc ctt cgg aga gct ctg tca cct
 L   F   H   W   A   S   R   H   S   P   F   S   I   L   R   R   A   L   S   P
cgc ctt gta gca ctt gtg tct ctc cca ctg gcc tcc act tct tgt ggg cac ttt ctg gat
 R   L   V   A   L   V   S   L   P   L   A   S   T   S   C   G   H   F   L   D
gcc cag gcc taa tcc agc gct ggc act cag tta att ctc tct tcc ttt aaa ggg aac aat
 A   Q   A   -   S   S   A   G   T   Q   L   I   L   S   S   F   K   G   N   N
tac cac ctt gag gaa gga ggc ctg aaa gct aag tct ttg cta ttg cat ccg cct tct tct
 Y   H   L   E   E   G   G   L   K   A   K   S   L   L   L   H   P   P   S   S
cac gaa gac ctg cag ctt gaa agg agg gga ggg aga aat agc cct aag ctc cag gaa gac
 H   E   D   L   Q   L   E   R   R   G   G   R   N   S   P   K   L   Q   E   D
caa gcc cag cct ctg tga tga ttc cga gga aat tca gga aat cct tgc taa tgg cca aac
 Q   A   Q   P   L   -   -   F   R   G   N   S   G   N   P   C   -   W   P   N
ttc ccg ggt ttg aga tgc aaa agg aaa tca gat cgc agg gta atg ctt tta cac taa gca
 F   P   G   L   R   C   K   R   K   S   D   R   R   V   M   L   L   H   -   A
aga aat tgc agg ata taa agc tgt gca gac cat tgc ctc tga tta aga aag cga ttc tga
 R   N   C   R   I   -   S   C   A   D   H   C   L   -   L   R   K   R   F   -
gat tcc agc ctg gaa gga acc ggg ccc cag tgt taa ctg tgg tta att ctg ggt gga ggg
 D   S   S   L   E   G   T   G   P   Q   C   -   L   W   L   I   L   G   G   G
gtg atg ggt gat aat ttc tcc ttt ttc ttt tct gta cta acc att aat act ggg ggg atg
 V   M   G   D   N   F   S   F   F   F   S   V   L   T   I   N   T   G   G   M
ggc agt taa aaa tca aag gaa gaa aac ata aag aaa gcc aaa tcc ttt tcc atc tgt agc
 G   S   -   K   S   K   E   E   N   I   K   K   A   K   S   F   S   I   C   S
cat tca agc tac tgc ctg gac acg ctc tcc ttg aag ctg ggg ctc ctc tct cag ccc cac
 H   S   S   Y   C   L   D   T   L   S   L   K   L   G   L   L   S   Q   P   H
cta aca tgg ccc acc tcc ttc ctg ggg tca ctc cgt gtt cct caa gca ctt caa act cat
 L   T   W   P   T   S   F   L   G   S   L   R   V   P   Q   A   L   Q   T   H
cct ctc ctg cct ctg agc tga gca tat ggg cct ctc ccc ttc ctt taa aag ctc cat ccg
 P   L   L   P   L   S   -   A   Y   G   P   L   P   F   L   -   K   L   H   P
tga ctg cct tct ggg ggt cag cca cat agc tga gtg agg acc aca tgc ttc aca cca aat
 -   L   P   S   G   G   Q   P   H   S   -   V   R   T   T   C   F   T   P   N
gat aca aga agc ttg ttc caa atc tat gtg ggt ttt ttg aca ttt tag aga gca gat gct
 D   T   R   S   L   F   Q   I   Y   V   G   F   L   T   F   -   R   A   D   A
ttc cca tgc ttt cta att ccc tat tac atc tat aca tct ctt tta aaa gat cct gca agt
 F   P   C   F   L   I   P   Y   Y   I   Y   T   S   L   L   K   D   P   A   S
cct ttt agt gtc tgg aca cag gtt att ccc tcc ctt tct ttt ttc cca att tca cct gcc
 P   F   S   V   W   T   Q   V   I   P   S   L   S   F   F   P   I   S   P   A
cca gtc ctc ctg gtc agt gct tct gtt tcc tgc aca agg aaa cag cct tgc aaa cat ggt
 P   V   L   L   V   S   A   S   V   S   C   T   R   K   Q   P   C   K   H   G
gaa ccc tga aag cat gct gga aat gaa aga agc cag gta caa aag aca aca tac tat atg
 E   P   -   K   H   A   G   N   E   R   S   Q   V   Q   K   T   T   Y   Y   M
att tca ttc aca aga aat gtg tgt aac aga aaa att tac aga gac aga aaa taa att cct
 I   S   F   T   R   N   V   C   N   R   K   I   Y   R   D   R   K   -   I   P
agt tgc tta ggg ctg gga gag tgg ggt gag gcc tct cta cag aga tgg caa tgc cca gga
 S   C   L   G   L   G   E   W   G   E   A   S   L   Q   R   W   Q   C   P   G
ttc aat ctg aag atg ttg ttt aga tat cac agc ata gtc gga caa tag tga gga cag tct
 F   N   L   K   M   L   F   R   Y   H   S   I   V   G   Q   -   -   G   Q   S
ggc att ggc gta cag atg aac aaa tag att tgt gca ata gaa gaa gcc aaa cca gat tcc
 G   I   G   V   Q   M   N   K   -   I   C   A   I   E   E   A   K   P   D   S
agc cta cct cca gtc acc tga ttt aac aaa gtc aaa gtc aca aat caa tag gga gag gat
 S   L   P   P   V   T   -   F   N   K   V   K   V   T   N   Q   -   G   E   D
ggt ctt ttc agt aaa tga tac cgt atc aat tga ctc gtc ata tac aga aca aaa tga act
 G   L   F   S   K   -   Y   R   I   N   -   L   V   I   Y   R   T   K   -   T
ttg gtc cca tac aca cac aag tca taa aag ttc act caa agt gaa tga tag acc tac att
 L   V   P   Y   T   H   K   S   -   K   F   T   Q   S   E   -   -   T   Y   I
tga aaa caa aag caa tgc agg atc cag gag aaa ggg tcc agg cag gag aat att ttt atg
 -   K   Q   K   Q   C   R   I   Q   E   K   G   S   R   Q   E   N   I   F   M
acc ttg gac taa gca aga gat tca taa aca gga cac aaa acc act aac cac caa aaa aaa
 T   L   D   -   A   R   D   S   -   T   G   H   K   T   T   N   H   Q   K   K
gga ctg ata aac atg aca att aaa atg aag aat tta tgt tta tca aaa aca aaa agg gga
 G   L   I   N   M   T   I   K   M   K   N   L   C   L   S   K   T   K   R   G
acc atc gac tgg gag aac ata gtt aca aca ctt gta tcc aac tga gaa ttt ata ttc agg
 T   I   D   W   E   N   I   V   T   T   L   V   S   N   -   E   F   I   F   R
tta gaa gac tca aaa aca aaa agg caa gca atc cac taa gaa caa att ctg ggc aga aga
 L   E   D   S   K   T   K   R   Q   A   I   H   -   E   Q   I   L   G   R   R
ctt cac aaa aga tct gca aat gac caa tga cat cag gta aac atg gat tga aac aac aag
 L   H   K   R   S   A   N   D   Q   -   H   Q   V   N   M   D   -   N   N   K
aag aca cac cca caa gat ccg tga caa ttt tga aaa tga aac cct gag gca ggc aga att
 K   T   H   P   Q   D   P   -   Q   F   -   K   -   N   P   E   A   G   R   I
cta agg tca tgt ccc aga ttc ccc ctc ttg gtg tcc gag ccc tgc aca gtc cac ttg agt
 L   R   S   C   P   R   F   P   L   L   V   S   E   P   C   T   V   H   L   S
agg ggc cag acc tgt gga tat gat gga tgt ctc ttc tgt gat cag gtt aca tca tat gcc
 R   G   Q   T   C   G   Y   D   G   C   L   F   C   D   Q   V   T   S   Y   A
aaa gat gga ggg att ttg tga tta agt ctc taa tca gtt tac ctt caa gtt cat caa aag
 K   D   G   G   I   L   -   L   S   L   -   S   V   Y   L   Q   V   H   Q   K
cga gag tgg cct agg ttg gcc tgg ctt act cag gtg agg cct tta aaa gag agc tca agc
 R   E   W   P   R   L   A   W   L   T   Q   V   R   P   L   K   E   S   S   S
ctg ccc tga gct tgg aga gat ttt cct gct agc ctt gaa gaa aca aac agc tgt gag ttc
 L   P   -   A   W   R   D   F   P   A   S   L   E   E   T   N   S   C   E   F
tgc agc ttc aca gaa atg aat ttg gcc gac aat cat gtg agc tcc att aga aat gca ccc
 C   S   F   T   E   M   N   L   A   D   N   H   V   S   S   I   R   N   A   P
taa gct ctc tca acc tct ccc atc ccc cac agc acc tat tta ctg att atc tgt aca tga
 -   A   L   S   T   S   P   I   P   H   S   T   Y   L   L   I   I   C   T   -
tag cag ggc agg aac tag act ctc agc ccc ctg agg aca gac ctt gtc tgt cag tgc att
 -   Q   G   R   N   -   T   L   S   P   L   R   T   D   L   V   C   Q   C   I
tca ctg tct gtc cca gca cct tgc aca gag cct ggc aca tat caa gaa cac ggc aca ttt
 S   L   S   V   P   A   P   C   T   E   P   G   T   Y   Q   E   H   G   T   F
tta ttg aga cag act cct caa agc tgg aat cta aaa cca atg gca cct aat ctc agc caa
 L   L   R   Q   T   P   Q   S   W   N   L   K   P   M   A   P   N   L   S   Q
aaa ggt taa tgt gac acc tgt gct ttt att acc cat ttc tgc aga tgg aag ggt tgg ttg  acc>atc, Thr>Ile
 K   G   -   C   D   T   C   A   F   I   T   H   F   C   R   W   K   G   W   L
ttt gtg gct atc tca agt gaa atg cag aat tag tga ata aaa ttt tca tct gaa ttt aaa
 F   V   A   I   S   S   E   M   Q   N   -   -   I   K   F   S   S   E   F   K
gaa aat ctt aa
 E   N   L
```

**5'3' Frame 2**

```
gggc gcg ccc ggg cac ggc ggt gct ggc ggg gcg cgg gcg ggg cag cgc aca ggc cgg ccc
  G   A   P   G   H   G   G   A   G   G   A   R   A   G   Q   R   T   G   R   P
tct cgc ggg act cag ttt ccc cgt ctg cgc gcc cca cat att cgc ttt ccg cag gtt ctc
 S   R   G   T   Q   F   P   R   L   R   A   P   H   I   R   F   P   Q   V   L
tcg ctg aga ttc gcg tcc ttc aaa cta atg gaa ctt tgc aag ggg gcc ctg cgg cgg ctt
 S   L   R   F   A   S   F   K   L   M   E   L   C   K   G   A   L   R   R   L
cgc gtg gga gcg ccg ggg aaa gtt cct gca aat cgc cgg act ggg ggc ccg ccc ggg aag
 R   V   G   A   P   G   K   V   P   A   N   R   R   T   G   G   P   P   G   K
ctc gga ctg ggc cgg gca ggg act gca ggg gcc tct gcg cag acc cgg ggc gtt gcg gca
 L   G   L   G   R   A   G   T   A   G   A   S   A   Q   T   R   G   V   A   A
ccg cgg acc ccg gtc acg cgc tgg ttc tgc ccc cgg tac ccc gca cgc ggg acc ctg acc
 P   R   T   P   V   T   R   W   F   C   P   R   Y   P   A   R   G   T   L   T
gcg ggg cct ctg cgg ccg gac gaa ggc agc gtc cgc gga gct ggc aga aag ccc gcg ccc
 A   G   P   L   R   P   D   E   G   S   V   R   G   A   G   R   K   P   A   P
aga ccc aca gtg aag tga taa aag cag aga gca gaa aaa ctg agg gcc aga aag ggt gac
 R   P   T   V   K   -   -   K   Q   R   A   E   K   L   R   A   R   K   G   D
tcc cct gag tgt cat gcc aca agt tge agt gct gga gcc agg aca cct tcc aaa ccg gct
 S   P   E   C   H   A   T   S   X   S   A   G   A   R   T   P   S   K   P   A
gtg cct gcc tgc aag ccc cat cct cct cca tcc ccc tga gca gcc aga ctg cag agg gaa
 V   P   A   C   K   P   H   P   P   P   S   P   -   A   A   R   L   Q   R   E
tga gcg gac cga aaa cag tgt ggc cgc aac ttc ccc act gct ttg cgg gag gtg gag gga
 -   A   D   R   K   Q   C   G   R   N   F   P   T   A   L   R   E   V   E   G
gtt tct agg ctc ttt ccc tgt tgg gga att aag tag ctg ggg atc cca ctg agt ctc cac
 V   S   R   L   F   P   C   W   G   I   K   -   L   G   I   P   L   S   L   H
cac cct ctc acc cat ttt cca ccc ctc ccc cac cac ata tac att ctg ctc ccc cag tgc
 H   P   L   T   H   F   P   P   L   P   H   H   I   Y   I   L   L   P   Q   C
caa ata act tgc att ttc tgg aac aca cct gtc tgc tta cgg tct ctg tgc ctt tct ttg
 Q   I   T   C   I   F   W   N   T   P   V   C   L   R   S   L   C   L   S   L
ccc aca tct ccc ctc ttc ttg aaa cac ccc tcc tcc cct cca gcg tgg gct cat gtg acc
 P   T   S   P   L   F   L   K   H   P   S   S   P   P   A   W   A   H   V   T
tag aga agc acg tgg gag ccc tgt cat ctg ggg gac ttg ggt tca aat ccc act tct ggt
 -   R   S   T   W   E   P   C   H   L   G   D   L   G   S   N   P   T   S   G
cct taa gtc tgt gac tct ggg cct ttg ttt aat aac tgg agg agt ttc att agg aaa atg
 P   -   V   C   D   S   G   P   L   F   N   N   W   R   S   F   I   R   K   M
gca gtg acc aca tat gtc cct gga gtg act gat gtg cca ccc aca agc ttc tat ttg gtc
 A   V   T   T   Y   V   P   G   V   T   D   V   P   P   T   S   F   Y   L   V
agt gct ggg tcc cat cgt ccc atc tag cat gct cct ctt cca tcc tcg tgc ccc cac tca
 S   A   G   S   H   R   P   I   -   H   A   P   L   P   S   S   C   P   H   S
gtg cca ttg tct ctg cct ctc acc ctc tgt gct gct att cct gcc ctc atc acc tcc tcc
 V   P   L   S   L   P   L   T   L   C   A   A   I   P   A   L   I   T   S   S
cta ggg cag agg ccc cgt ctt ctt tac cat gcc cca gtg cca gca cag tgc ttt ggc aag
 L   G   Q   R   P   R   L   L   Y   H   A   P   V   P   A   Q   C   F   G   K
tta ttc agc act ggg gga gta gag tgt atg tgt ggt ggg gga ggg ggt gga gaa tgg gta
 L   F   S   T   G   G   V   E   C   M   C   G   G   G   G   G   G   E   W   V
aga ggg tga tgg aga ctc agg ggg acc ccc agc tac tta ata ccc ctg agg aag tgt cgg
 R   G   -   W   R   L   R   G   T   P   S   Y   L   I   P   L   R   K   C   R
ccg aag tgg acc cac tcc tgt gac agt tca cag cct aga aga ggc gtc tgt aac cac ggt
 P   K   W   T   H   S   C   D   S   S   Q   P   R   R   G   V   C   N   H   G
gac cag taa gtg cca gcc ctg ggg cca cgt ggc tgt tct aga gaa gga aga cag cac tac
 D   Q   -   V   P   A   L   G   P   R   G   C   S   R   E   G   R   Q   H   Y
cct gag agc cat gct ggt ggc ttc atc agg aag gga gtc agc agg ggg agg act ggg gga
 P   E   S   H   A   G   G   F   I   R   K   G   V   S   R   G   R   T   G   G
cca agc ggt ccc taa gaa gca gaa ttt gca ata ccc aag gtt tta gaa aga gca acc tgg
 P   S   G   P   -   E   A   E   F   A   I   P   K   V   L   E   R   A   T   W
cag ggc cac tgc aca atg aga tca cag act cac ctc cca gct gtg agt gca gct cac ccc
 Q   G   H   C   T   M   R   S   Q   T   H   L   P   A   V   S   A   A   H   P
acc tgc tct gac cca ggc acc ggt tat gaa gaa gac aag cag ctc tac aga ggg gcc tca
 T   C   S   D   P   G   T   G   Y   E   E   D   K   Q   L   Y   R   G   A   S
aga aat cct tcg aaa cag aaa tca aag tct tgc tgg aag ggt tct ggg cac cct gga ctg
 R   N   P   S   K   Q   K   S   K   S   C   W   K   G   S   G   H   P   G   L
tcc aca tgg gag gct gtt ggt gcg tgt gcc tac gtg tgt cca tca gtg gat gca cga aga
 S   T   W   E   A   V   G   A   C   A   Y   V   C   P   S   V   D   A   R   R
agg aaa tga att gat tcc att ctt gat gtg act ttc agc tgt eca gct gtc tgc act ccc
 R   K   -   I   D   S   I   L   D   V   T   F   S   C   X   A   V   C   T   P
aga cgg ggt gat ggc ctg gca cca acg gag cat tac cct gat gaa tca tgg tcg agg tac
 R   R   G   D   G   L   A   P   T   E   H   Y   P   D   E   S   W   S   R   Y
ttt tcc act ggg cca gcc gcc aca gcc cct tct cca tcc ttc gga gag ctc tgt cac ctc
 F   S   T   G   P   A   A   T   A   P   S   P   S   F   G   E   L   C   H   L
gcc ttg tag cac ttg tgt ctc tcc cac tgg cct cca ctt ctt gtg ggc act ttc tgg atg
 A   L   -   H   L   C   L   S   H   W   P   P   L   L   V   G   T   F   W   M
ccc agg cct aat cca gcg ctg gca ctc agt taa ttc tct ctt cct tta aag gga aca att
 P   R   P   N   P   A   L   A   L   S   -   F   S   L   P   L   K   G   T   I
acc acc ttg agg aag gag gcc tga aag cta agt ctt tgc tat tgc atc cgc ctt ctt ctc
 T   T   L   R   K   E   A   -   K   L   S   L   C   Y   C   I   R   L   L   L
acg aag acc tgc agc ttg aaa gga ggg gag gga gaa ata gcc cta agc tcc agg aag acc
 T   K   T   C   S   L   K   G   G   E   G   E   I   A   L   S   S   R   K   T
aag ccc agc ctc tgt gat gat tcc gag gaa att cag gaa atc ctt gct aat ggc caa act
 K   P   S   L   C   D   D   S   E   E   I   Q   E   I   L   A   N   G   Q   T
tcc cgg gtt tga gat gca aaa gga aat cag atc gca ggg taa tgc ttt tac act aag caa
 S   R   V   -   D   A   K   G   N   Q   I   A   G   -   C   F   Y   T   K   Q
gaa att gca gga tat aaa gct gtg cag acc att gcc tct gat taa gaa agc gat tct gag
 E   I   A   G   Y   K   A   V   Q   T   I   A   S   D   -   E   S   D   S   E
att cca gcc tgg aag gaa ccg ggc ccc agt gtt aac tgt ggt taa ttc tgg gtg gag ggg
 I   P   A   W   K   E   P   G   P   S   V   N   C   G   -   F   W   V   E   G
tga tgg gtg ata att tct cct ttt tct ttt ctg tac taa cca tta ata ctg ggg gga tgg
 -   W   V   I   I   S   P   F   S   F   L   Y   -   P   L   I   L   G   G   W
gca gtt aaa aat caa agg aag aaa aca taa aga aag cca aat cct ttt cca tct gta gcc
 A   V   K   N   Q   R   K   K   T   -   R   K   P   N   P   F   P   S   V   A
att caa gct act gcc tgg aca cgc tct cct tga agc tgg ggc tcc tct ctc agc ccc acc
 I   Q   A   T   A   W   T   R   S   P   -   S   W   G   S   S   L   S   P   T
taa cat ggc cca cct cct tcc tgg ggt cac tcc gtg ttc ctc aag cac ttc aaa ctc atc
 -   H   G   P   P   P   S   W   G   H   S   V   F   L   K   H   F   K   L   I
ctc tcc tgc ctc tga gct gag cat atg ggc ctc tcc cct tcc ttt aaa agc tcc atc cgt
 L   S   C   L   -   A   E   H   M   G   L   S   P   S   F   K   S   S   I   R
gac tgc ctt ctg ggg gtc agc cac ata gct gag tga gga cca cat gct tca cac caa atg
 D   C   L   L   G   V   S   H   I   A   E   -   G   P   H   A   S   H   Q   M
ata caa gaa gct tgt tcc aaa tct atg tgg gtt ttt tga cat ttt aga gag cag atg ctt
 I   Q   E   A   C   S   K   S   M   W   V   F   -   H   F   R   E   Q   M   L
tcc cat gct ttc taa ttc cct att aca tct ata cat ctc ttt taa aag atc ctg caa gtc
 S   H   A   F   -   F   P   I   T   S   I   H   L   F   -   K   I   L   Q   V
ctt tta gtg tct gga cac agg tta ttc cct ccc ttt ctt ttt tcc caa ttt cac ctg ccc
 L   L   V   S   G   H   R   L   F   P   P   F   L   F   S   Q   F   H   L   P
cag tcc tcc tgg tca gtg ctt ctg ttt cct gca caa gga aac agc ctt gca aac atg gtg
 Q   S   S   W   S   V   L   L   F   P   A   Q   G   N   S   L   A   N   M   V
aac cct gaa agc atg ctg gaa atg aaa gaa gcc agg tac aaa aga caa cat act ata tga
 N   P   E   S   M   L   E   M   K   E   A   R   Y   K   R   Q   H   T   I   -
ttt cat tca caa gaa atg tgt gta aca gaa aaa ttt aca gag aca gaa aat aaa ttc cta
 F   H   S   Q   E   M   C   V   T   E   K   F   T   E   T   E   N   K   F   L
gtt gct tag ggc tgg gag agt ggg gtg agg cct ctc tac aga gat ggc aat gcc cag gat
 V   A   -   G   W   E   S   G   V   R   P   L   Y   R   D   G   N   A   Q   D
tca atc tga aga tgt tgt tta gat atc aca gca tag tcg gac aat agt gag gac agt ctg
 S   I   -   R   C   C   L   D   I   T   A   -   S   D   N   S   E   D   S   L
gca ttg gcg tac aga tga aca aat aga ttt gtg caa tag aag aag cca aac cag att cca
 A   L   A   Y   R   -   T   N   R   F   V   Q   -   K   K   P   N   Q   I   P
gcc tac ctc cag tca cct gat tta aca aag tca aag tca caa atc aat agg gag agg atg
 A   Y   L   Q   S   P   D   L   T   K   S   K   S   Q   I   N   R   E   R   M
gtc ttt tca gta aat gat acc gta tca att gac tcg tca tat aca gaa caa aat gaa ctt
 V   F   S   V   N   D   T   V   S   I   D   S   S   Y   T   E   Q   N   E   L
tgg tcc cat aca cac aca agt cat aaa agt tca ctc aaa gtg aat gat aga cct aca ttt
 W   S   H   T   H   T   S   H   K   S   S   L   K   V   N   D   R   P   T   F
gaa aac aaa agc aat gca gga tcc agg aga aag ggt cca ggc agg aga ata ttt tta tga
 E   N   K   S   N   A   G   S   R   R   K   G   P   G   R   R   I   F   L   -
cct tgg act aag caa gag att cat aaa cag gac aca aaa cca cta acc acc aaa aaa aag
 P   W   T   K   Q   E   I   H   K   Q   D   T   K   P   L   T   T   K   K   K
gac tga taa aca tga caa tta aaa tga aga att tat gtt tat caa aaa caa aaa ggg gaa
 D   -   -   T   -   Q   L   K   -   R   I   Y   V   Y   Q   K   Q   K   G   E
cca tcg act ggg aga aca tag tta caa cac ttg tat cca act gag aat tta tat tca ggt
 P   S   T   G   R   T   -   L   Q   H   L   Y   P   T   E   N   L   Y   S   G
tag aag act caa aaa caa aaa ggc aag caa tcc act aag aac aaa ttc tgg gca gaa gac
 -   K   T   Q   K   Q   K   G   K   Q   S   T   K   N   K   F   W   A   E   D
ttc aca aaa gat ctg caa atg acc aat gac atc agg taa aca tgg att gaa aca aca aga
 F   T   K   D   L   Q   M   T   N   D   I   R   -   T   W   I   E   T   T   R
aga cac acc cac aag atc cgt gac aat ttt gaa aat gaa acc ctg agg cag gca gaa ttc
 R   H   T   H   K   I   R   D   N   F   E   N   E   T   L   R   Q   A   E   F
taa ggt cat gtc cca gat tcc ccc tct tgg tgt ccg agc cct gca cag tcc act tga gta
 -   G   H   V   P   D   S   P   S   W   C   P   S   P   A   Q   S   T   -   V
ggg gcc aga cct gtg gat atg atg gat gtc tct tct gtg atc agg tta cat cat atg cca
 G   A   R   P   V   D   M   M   D   V   S   S   V   I   R   L   H   H   M   P
aag atg gag gga ttt tgt gat taa gtc tct aat cag ttt acc ttc aag ttc atc aaa agc
 K   M   E   G   F   C   D   -   V   S   N   Q   F   T   F   K   F   I   K   S
gag agt ggc cta ggt tgg cct ggc tta ctc agg tga ggc ctt taa aag aga gct caa gcc
 E   S   G   L   G   W   P   G   L   L   R   -   G   L   -   K   R   A   Q   A
tgc cct gag ctt gga gag att ttc ctg cta gcc ttg aag aaa caa aca gct gtg agt tct
 C   P   E   L   G   E   I   F   L   L   A   L   K   K   Q   T   A   V   S   S
gca gct tca cag aaa tga att tgg ccg aca atc atg tga gct cca tta gaa atg cac cct
 A   A   S   Q   K   -   I   W   P   T   I   M   -   A   P   L   E   M   H   P
aag ctc tct caa cct ctc cca tcc ccc aca gca cct att tac tga tta tct gta cat gat
 K   L   S   Q   P   L   P   S   P   T   A   P   I   Y   -   L   S   V   H   D
agc agg gca gga act aga ctc tca gcc ccc tga gga cag acc ttg tct gtc agt gca ttt
 S   R   A   G   T   R   L   S   A   P   -   G   Q   T   L   S   V   S   A   F
cac tgt ctg tcc cag cac ctt gca cag agc ctg gca cat atc aag aac acg gca cat ttt
 H   C   L   S   Q   H   L   A   Q   S   L   A   H   I   K   N   T   A   H   F
tat tga gac aga ctc ctc aaa gct gga atc taa aac caa tgg cac cta atc tca gcc aaa
 Y   -   D   R   L   L   K   A   G   I   -   N   Q   W   H   L   I   S   A   K
aag gtt aat gtg aca cct gtg ctt tta tta ccc att tct gca gat gga agg gtt ggt tgt
 K   V   N   V   T   P   V   L   L   L   P   I   S   A   D   G   R   V   G   C
ttg tgg cta tct caa gtg aaa tgc aga att agt gaa taa aat ttt cat ctg aat tta aag
 L   W   L   S   Q   V   K   C   R   I   S   E   -   N   F   H   L   N   L   K
aaa atc tta a
 K   I   L
```

**5'3' Frame 3**

```
gggcg cgc ccg ggc acg gcg gtg ctg gcg ggg cgc ggg cgg ggc agc gca cag gcc ggc cct
   A   R   P   G   T   A   V   L   A   G   R   G   R   G   S   A   Q   A   G   P
ctc gcg gga ctc agt ttc ccc gtc tgc gcg ccc cac ata ttc gct ttc cgc agg ttc tct
 L   A   G   L   S   F   P   V   C   A   P   H   I   F   A   F   R   R   F   S
cgc tga gat tcg cgt cct tca aac taa tgg aac ttt gca agg ggg ccc tgc ggc ggc ttc
 R   -   D   S   R   P   S   N   -   W   N   F   A   R   G   P   C   G   G   F
gcg tgg gag cgc cgg gga aag ttc ctg caa atc gcc gga ctg ggg gcc cgc ccg gga agc
 A   W   E   R   R   G   K   F   L   Q   I   A   G   L   G   A   R   P   G   S
tcg gac tgg gcc ggg cag gga ctg cag ggg cct ctg cgc aga ccc ggg gcg ttg cgg cac
 S   D   W   A   G   Q   G   L   Q   G   P   L   R   R   P   G   A   L   R   H
cgc gga ccc cgg tca cgc gct ggt tct gcc ccc ggt acc ccg cac gcg gga ccc tga ccg
 R   G   P   R   S   R   A   G   S   A   P   G   T   P   H   A   G   P   -   P
cgg ggc ctc tgc ggc cgg acg aag gca gcg tcc gcg gag ctg gca gaa agc ccg cgc cca
 R   G   L   C   G   R   T   K   A   A   S   A   E   L   A   E   S   P   R   P
gac cca cag tga agt gat aaa agc aga gag cag aaa aac tga ggg cca gaa agg gtg act
 D   P   Q   -   S   D   K   S   R   E   Q   K   N   -   G   P   E   R   V   T
ccc ctg agt gtc atg cca caa gtt gea gtg ctg gag cca gga cac ctt cca aac cgg ctg
 P   L   S   V   M   P   Q   V   X   V   L   E   P   G   H   L   P   N   R   L
tgc ctg cct gca agc ccc atc ctc ctc cat ccc cct gag cag cca gac tgc aga ggg aat
 C   L   P   A   S   P   I   L   L   H   P   P   E   Q   P   D   C   R   G   N
gag cgg acc gaa aac agt gtg gcc gca act tcc cca ctg ctt tgc ggg agg tgg agg gag
 E   R   T   E   N   S   V   A   A   T   S   P   L   L   C   G   R   W   R   E
ttt cta ggc tct ttc cct gtt ggg gaa tta agt agc tgg gga tcc cac tga gtc tcc acc
 F   L   G   S   F   P   V   G   E   L   S   S   W   G   S   H   -   V   S   T
acc ctc tca ccc att ttc cac ccc tcc ccc acc aca tat aca ttc tgc tcc ccc agt gcc
 T   L   S   P   I   F   H   P   S   P   T   T   Y   T   F   C   S   P   S   A
aaa taa ctt gca ttt tct gga aca cac ctg tct gct tac ggt ctc tgt gcc ttt ctt tgc
 K   -   L   A   F   S   G   T   H   L   S   A   Y   G   L   C   A   F   L   C
cca cat ctc ccc tct tct tga aac acc cct cct ccc ctc cag cgt ggg ctc atg tga cct
 P   H   L   P   S   S   -   N   T   P   P   P   L   Q   R   G   L   M   -   P
aga gaa gca cgt ggg agc cct gtc atc tgg ggg act tgg gtt caa atc cca ctt ctg gtc
 R   E   A   R   G   S   P   V   I   W   G   T   W   V   Q   I   P   L   L   V
ctt aag tct gtg act ctg ggc ctt tgt tta ata act gga gga gtt tca tta gga aaa tgg
 L   K   S   V   T   L   G   L   C   L   I   T   G   G   V   S   L   G   K   W
cag tga cca cat atg tcc ctg gag tga ctg atg tgc cac cca caa gct tct att tgg tca
 Q   -   P   H   M   S   L   E   -   L   M   C   H   P   Q   A   S   I   W   S
gtg ctg ggt ccc atc gtc cca tct agc atg ctc ctc ttc cat cct cgt gcc ccc act cag
 V   L   G   P   I   V   P   S   S   M   L   L   F   H   P   R   A   P   T   Q
tgc cat tgt ctc tgc ctc tca ccc tct gtg ctg cta ttc ctg ccc tca tca cct cct ccc
 C   H   C   L   C   L   S   P   S   V   L   L   F   L   P   S   S   P   P   P
tag ggc aga ggc ccc gtc ttc ttt acc atg ccc cag tgc cag cac agt gct ttg gca agt
 -   G   R   G   P   V   F   F   T   M   P   Q   C   Q   H   S   A   L   A   S
tat tca gca ctg ggg gag tag agt gta tgt gtg gtg ggg gag ggg gtg gag aat ggg taa
 Y   S   A   L   G   E   -   S   V   C   V   V   G   E   G   V   E   N   G   -
gag ggt gat gga gac tca ggg gga ccc cca gct act taa tac ccc tga gga agt gtc ggc
 E   G   D   G   D   S   G   G   P   P   A   T   -   Y   P   -   G   S   V   G
cga agt gga ccc act cct gtg aca gtt cac agc cta gaa gag gcg tct gta acc acg gtg
 R   S   G   P   T   P   V   T   V   H   S   L   E   E   A   S   V   T   T   V
acc agt aag tgc cag ccc tgg ggc cac gtg gct gtt cta gag aag gaa gac agc act acc
 T   S   K   C   Q   P   W   G   H   V   A   V   L   E   K   E   D   S   T   T
ctg aga gcc atg ctg gtg gct tca tca gga agg gag tca gca ggg gga gga ctg ggg gac
 L   R   A   M   L   V   A   S   S   G   R   E   S   A   G   G   G   L   G   D
caa gcg gtc cct aag aag cag aat ttg caa tac cca agg ttt tag aaa gag caa cct ggc
 Q   A   V   P   K   K   Q   N   L   Q   Y   P   R   F   -   K   E   Q   P   G
agg gcc act gca caa tga gat cac aga ctc acc tcc cag ctg tga gtg cag ctc acc cca
 R   A   T   A   Q   -   D   H   R   L   T   S   Q   L   -   V   Q   L   T   P
cct gct ctg acc cag gca ccg gtt atg aag aag aca agc agc tct aca gag ggg cct caa
 P   A   L   T   Q   A   P   V   M   K   K   T   S   S   S   T   E   G   P   Q
gaa atc ctt cga aac aga aat caa agt ctt gct gga agg gtt ctg ggc acc ctg gac tgt
 E   I   L   R   N   R   N   Q   S   L   A   G   R   V   L   G   T   L   D   C
cca cat ggg agg ctg ttg gtg cgt gtg cct acg tgt gtc cat cag tgg atg cac gaa gaa
 P   H   G   R   L   L   V   R   V   P   T   C   V   H   Q   W   M   H   E   E
gga aat gaa ttg att cca ttc ttg atg tga ctt tca gct gte cag ctg tct gca ctc cca
 G   N   E   L   I   P   F   L   M   -   L   S   A   X   Q   L   S   A   L   P
gac ggg gtg atg gcc tgg cac caa cgg agc att acc ctg atg aat cat ggt cga ggt act  SNP G/A=C/T, Thr>Ile
 D   G   V   M   A   W   H   Q   R   S   I   T   L   M   N   H   G   R   G   T
ttt cca ctg ggc cag ccg cca cag ccc ctt ctc cat cct tcg gag agc tct gtc acc tcg
 F   P   L   G   Q   P   P   Q   P   L   L   H   P   S   E   S   S   V   T   S
cct tgt agc act tgt gtc tct ccc act ggc ctc cac ttc ttg tgg gca ctt tct gga tgc
 P   C   S   T   C   V   S   P   T   G   L   H   F   L   W   A   L   S   G   C
cca ggc cta atc cag cgc tgg cac tca gtt aat tct ctc ttc ctt taa agg gaa caa tta
 P   G   L   I   Q   R   W   H   S   V   N   S   L   F   L   -   R   E   Q   L
cca cct tga gga agg agg cct gaa agc taa gtc ttt gct att gca tcc gcc ttc ttc tca
 P   P   -   G   R   R   P   E   S   -   V   F   A   I   A   S   A   F   F   S
cga aga cct gca gct tga aag gag ggg agg gag aaa tag ccc taa gct cca gga aga cca
 R   R   P   A   A   -   K   E   G   R   E   K   -   P   -   A   P   G   R   P
agc cca gcc tct gtg atg att ccg agg aaa ttc agg aaa tcc ttg cta atg gcc aaa ctt
 S   P   A   S   V   M   I   P   R   K   F   R   K   S   L   L   M   A   K   L
ccc ggg ttt gag atg caa aag gaa atc aga tcg cag ggt aat gct ttt aca cta agc aag
 P   G   F   E   M   Q   K   E   I   R   S   Q   G   N   A   F   T   L   S   K
aaa ttg cag gat ata aag ctg tgc aga cca ttg cct ctg att aag aaa gcg att ctg aga
 K   L   Q   D   I   K   L   C   R   P   L   P   L   I   K   K   A   I   L   R
ttc cag cct gga agg aac cgg gcc cca gtg tta act gtg gtt aat tct ggg tgg agg ggt
 F   Q   P   G   R   N   R   A   P   V   L   T   V   V   N   S   G   W   R   G
gat ggg tga taa ttt ctc ctt ttt ctt ttc tgt act aac cat taa tac tgg ggg gat ggg
 D   G   -   -   F   L   L   F   L   F   C   T   N   H   -   Y   W   G   D   G
cag tta aaa atc aaa gga aga aaa cat aaa gaa agc caa atc ctt ttc cat ctg tag cca
 Q   L   K   I   K   G   R   K   H   K   E   S   Q   I   L   F   H   L   -   P
ttc aag cta ctg cct gga cac gct ctc ctt gaa gct ggg gct cct ctc tca gcc cca cct
 F   K   L   L   P   G   H   A   L   L   E   A   G   A   P   L   S   A   P   P
aac atg gcc cac ctc ctt cct ggg gtc act ccg tgt tcc tca agc act tca aac tca tcc
 N   M   A   H   L   L   P   G   V   T   P   C   S   S   S   T   S   N   S   S
tct cct gcc tct gag ctg agc ata tgg gcc tct ccc ctt cct tta aaa gct cca tcc gtg
 S   P   A   S   E   L   S   I   W   A   S   P   L   P   L   K   A   P   S   V
act gcc ttc tgg ggg tca gcc aca tag ctg agt gag gac cac atg ctt cac acc aaa tga
 T   A   F   W   G   S   A   T   -   L   S   E   D   H   M   L   H   T   K   -
tac aag aag ctt gtt cca aat cta tgt ggg ttt ttt gac att tta gag agc aga tgc ttt
 Y   K   K   L   V   P   N   L   C   G   F   F   D   I   L   E   S   R   C   F
ccc atg ctt tct aat tcc cta tta cat cta tac atc tct ttt aaa aga tcc tgc aag tcc
 P   M   L   S   N   S   L   L   H   L   Y   I   S   F   K   R   S   C   K   S
ttt tag tgt ctg gac aca ggt tat tcc ctc cct ttc ttt ttt ccc aat ttc acc tgc ccc
 F   -   C   L   D   T   G   Y   S   L   P   F   F   F   P   N   F   T   C   P
agt cct cct ggt cag tgc ttc tgt ttc ctg cac aag gaa aca gcc ttg caa aca tgg tga
 S   P   P   G   Q   C   F   C   F   L   H   K   E   T   A   L   Q   T   W   -
acc ctg aaa gca tgc tgg aaa tga aag aag cca ggt aca aaa gac aac ata cta tat gat
 T   L   K   A   C   W   K   -   K   K   P   G   T   K   D   N   I   L   Y   D
ttc att cac aag aaa tgt gtg taa cag aaa aat tta cag aga cag aaa ata aat tcc tag
 F   I   H   K   K   C   V   -   Q   K   N   L   Q   R   Q   K   I   N   S   -
ttg ctt agg gct ggg aga gtg ggg tga ggc ctc tct aca gag atg gca atg ccc agg att
 L   L   R   A   G   R   V   G   -   G   L   S   T   E   M   A   M   P   R   I
caa tct gaa gat gtt gtt tag ata tca cag cat agt cgg aca ata gtg agg aca gtc tgg
 Q   S   E   D   V   V   -   I   S   Q   H   S   R   T   I   V   R   T   V   W
cat tgg cgt aca gat gaa caa ata gat ttg tgc aat aga aga agc caa acc aga ttc cag
 H   W   R   T   D   E   Q   I   D   L   C   N   R   R   S   Q   T   R   F   Q
cct acc tcc agt cac ctg att taa caa agt caa agt cac aaa tca ata ggg aga gga tgg
 P   T   S   S   H   L   I   -   Q   S   Q   S   H   K   S   I   G   R   G   W
tct ttt cag taa atg ata ccg tat caa ttg act cgt cat ata cag aac aaa atg aac ttt
 S   F   Q   -   M   I   P   Y   Q   L   T   R   H   I   Q   N   K   M   N   F
ggt ccc ata cac aca caa gtc ata aaa gtt cac tca aag tga atg ata gac cta cat ttg
 G   P   I   H   T   Q   V   I   K   V   H   S   K   -   M   I   D   L   H   L
aaa aca aaa gca atg cag gat cca gga gaa agg gtc cag gca gga gaa tat ttt tat gac
 K   T   K   A   M   Q   D   P   G   E   R   V   Q   A   G   E   Y   F   Y   D
ctt gga cta agc aag aga ttc ata aac agg aca caa aac cac taa cca cca aaa aaa agg
 L   G   L   S   K   R   F   I   N   R   T   Q   N   H   -   P   P   K   K   R
act gat aaa cat gac aat taa aat gaa gaa ttt atg ttt atc aaa aac aaa aag ggg aac
 T   D   K   H   D   N   -   N   E   E   F   M   F   I   K   N   K   K   G   N
cat cga ctg gga gaa cat agt tac aac act tgt atc caa ctg aga att tat att cag gtt
 H   R   L   G   E   H   S   Y   N   T   C   I   Q   L   R   I   Y   I   Q   V
aga aga ctc aaa aac aaa aag gca agc aat cca cta aga aca aat tct ggg cag aag act
 R   R   L   K   N   K   K   A   S   N   P   L   R   T   N   S   G   Q   K   T
tca caa aag atc tgc aaa tga cca atg aca tca ggt aaa cat gga ttg aaa caa caa gaa
 S   Q   K   I   C   K   -   P   M   T   S   G   K   H   G   L   K   Q   Q   E
gac aca ccc aca aga tcc gtg aca att ttg aaa atg aaa ccc tga ggc agg cag aat tct
 D   T   P   T   R   S   V   T   I   L   K   M   K   P   -   G   R   Q   N   S
aag gtc atg tcc cag att ccc cct ctt ggt gtc cga gcc ctg cac agt cca ctt gag tag
 K   V   M   S   Q   I   P   P   L   G   V   R   A   L   H   S   P   L   E   -
ggg cca gac ctg tgg ata tga tgg atg tct ctt ctg tga tca ggt tac atc ata tgc caa
 G   P   D   L   W   I   -   W   M   S   L   L   -   S   G   Y   I   I   C   Q
aga tgg agg gat ttt gtg att aag tct cta atc agt tta cct tca agt tca tca aaa gcg
 R   W   R   D   F   V   I   K   S   L   I   S   L   P   S   S   S   S   K   A
aga gtg gcc tag gtt ggc ctg gct tac tca ggt gag gcc ttt aaa aga gag ctc aag cct
 R   V   A   -   V   G   L   A   Y   S   G   E   A   F   K   R   E   L   K   P
gcc ctg agc ttg gag aga ttt tcc tgc tag cct tga aga aac aaa cag ctg tga gtt ctg
 A   L   S   L   E   R   F   S   C   -   P   -   R   N   K   Q   L   -   V   L
cag ctt cac aga aat gaa ttt ggc cga caa tca tgt gag ctc cat tag aaa tgc acc cta
 Q   L   H   R   N   E   F   G   R   Q   S   C   E   L   H   -   K   C   T   L
agc tct ctc aac ctc tcc cat ccc cca cag cac cta ttt act gat tat ctg tac atg ata
 S   S   L   N   L   S   H   P   P   Q   H   L   F   T   D   Y   L   Y   M   I
gca ggg cag gaa cta gac tct cag ccc cct gag gac aga cct tgt ctg tca gtg cat ttc
 A   G   Q   E   L   D   S   Q   P   P   E   D   R   P   C   L   S   V   H   F
act gtc tgt ccc agc acc ttg cac aga gcc tgg cac ata tca aga aca cgg cac att ttt
 T   V   C   P   S   T   L   H   R   A   W   H   I   S   R   T   R   H   I   F
att gag aca gac tcc tca aag ctg gaa tct aaa acc aat ggc acc taa tct cag cca aaa
 I   E   T   D   S   S   K   L   E   S   K   T   N   G   T   -   S   Q   P   K
agg tta atg tga cac ctg tgc ttt tat tac cca ttt ctg cag atg gaa ggg ttg gtt gtt
 R   L   M   -   H   L   C   F   Y   Y   P   F   L   Q   M   E   G   L   V   V
tgt ggc tat ctc aag tga aat gca gaa tta gtg aat aaa att ttc atc tga att taa aga
 C   G   Y   L   K   -   N   A   E   L   V   N   K   I   F   I   -   I   -   R
aaa tct taa
 K   S   -
```

1) **LncRNA *ENST00000647969.1***

rs61871745

**5'3' Frame 1**

```
tct gac cca ggc acc ggt tat gaa gaa gac aag cag ctc tac aga ggg gcc tca aga aat
 S   D   P   G   T   G   Y   E   E   D   K   Q   L   Y   R   G   A   S   R   N
cct tcg aaa cag aaa tca aag tct tgc tgg aag ggt tct ggg cac cct gga ctg tcc aca
 P   S   K   Q   K   S   K   S   C   W   K   G   S   G   H   P   G   L   S   T
tgg gag gct gtt ggt gcg tgt gcc tac gtg tgt cca tca gtg gat gca cga aga agg aaa
 W   E   A   V   G   A   C   A   Y   V   C   P   S   V   D   A   R   R   R   K
tgc tgt cag ctg tct gca ctc cca gac ggg gtg atg gcc tgg cac caa cgg agc att acc  G/A=C/T, Thr > Ile
 C   C   Q   L   S   A   L   P   D   G   V   M   A   W   H   Q   R   S   I   T
ctg atg aat cat ggt cga ggt act ttt cca ctg ggc cag ccg cca cag ccc ctt ctc cat
 L   M   N   H   G   R   G   T   F   P   L   G   Q   P   P   Q   P   L   L   H
cct tcg gag agc tct gtc acc tcg cct tgt agc act tgt gtc tct ccc act ggc ctc cac
 P   S   E   S   S   V   T   S   P   C   S   T   C   V   S   P   T   G   L   H
ttc ttg tgg gca ctt tct gga tgc cca ggc cta atc cag cgc tgg cac tca gtt aat tct
 F   L   W   A   L   S   G   C   P   G   L   I   Q   R   W   H   S   V   N   S
ctc ttc ctt taa agg gaa caa tta cca cct tga gga agg agg cct gaa agc taa gtc ttt
 L   F   L   -   R   E   Q   L   P   P   -   G   R   R   P   E   S   -   V   F
gct att gca tcc gcc ttc ttc tca cga aga cct gca gct tga aag gag ggg agg gag aaa
 A   I   A   S   A   F   F   S   R   R   P   A   A   -   K   E   G   R   E   K
tag ccc taa gct cca gga aga cca
 -   P   -   A   P   G   R   P
```

**5'3' Frame 2**

```
tctg acc cag gca ccg gtt atg aag aag aca agc agc tct aca gag ggg cct caa gaa atc
  L   T   Q   A   P   V   M   K   K   T   S   S   S   T   E   G   P   Q   E   I
ctt cga aac aga aat caa agt ctt gct gga agg gtt ctg ggc acc ctg gac tgt cca cat
 L   R   N   R   N   Q   S   L   A   G   R   V   L   G   T   L   D   C   P   H
ggg agg ctg ttg gtg cgt gtg cct acg tgt gtc cat cag tgg atg cac gaa gaa gga aat
 G   R   L   L   V   R   V   P   T   C   V   H   Q   W   M   H   E   E   G   N
gct gtc agc tgt ctg cac tcc cag acg ggg tga tgg cct ggc acc aac gga gca tta ccc
 A   V   S   C   L   H   S   Q   T   G   -   W   P   G   T   N   G   A   L   P
tga tga atc atg gtc gag gta ctt ttc cac tgg gcc agc cgc cac agc ccc ttc tcc atc
 -   -   I   M   V   E   V   L   F   H   W   A   S   R   H   S   P   F   S   I
ctt cgg aga gct ctg tca cct cgc ctt gta gca ctt gtg tct ctc cca ctg gcc tcc act
 L   R   R   A   L   S   P   R   L   V   A   L   V   S   L   P   L   A   S   T
tct tgt ggg cac ttt ctg gat gcc cag gcc taa tcc agc gct ggc act cag tta att ctc
 S   C   G   H   F   L   D   A   Q   A   -   S   S   A   G   T   Q   L   I   L
tct tcc ttt aaa ggg aac aat tac cac ctt gag gaa gga ggc ctg aaa gct aag tct ttg
 S   S   F   K   G   N   N   Y   H   L   E   E   G   G   L   K   A   K   S   L
cta ttg cat ccg cct tct tct cac gaa gac ctg cag ctt gaa agg agg gga ggg aga aat
 L   L   H   P   P   S   S   H   E   D   L   Q   L   E   R   R   G   G   R   N
agc cct aag ctc cag gaa gac caa
 S   P   K   L   Q   E   D   Q
```

**5'3' Frame 3**

```
tctga ccc agg cac cgg tta tga aga aga caa gca gct cta cag agg ggc ctc aag aaa tcc
   -   P   R   H   R   L   -   R   R   Q   A   A   L   Q   R   G   L   K   K   S
ttc gaa aca gaa atc aaa gtc ttg ctg gaa ggg ttc tgg gca ccc tgg act gtc cac atg
 F   E   T   E   I   K   V   L   L   E   G   F   W   A   P   W   T   V   H   M
gga ggc tgt tgg tgc gtg tgc cta cgt gtg tcc atc agt gga tgc acg aag aag gaa atg
 G   G   C   W   C   V   C   L   R   V   S   I   S   G   C   T   K   K   E   M
ctg tca gct gtc tgc act ccc aga cgg ggt gat ggc ctg gca cca acg gag cat tac cct synonymous mutation
 L   S   A   V   C   T   P   R   R   G   D   G   L   A   P   T   E   H   Y   P
gat gaa tca tgg tcg agg tac ttt tcc act ggg cca gcc gcc aca gcc cct tct cca tcc
 D   E   S   W   S   R   Y   F   S   T   G   P   A   A   T   A   P   S   P   S
ttc gga gag ctc tgt cac ctc gcc ttg tag cac ttg tgt ctc tcc cac tgg cct cca ctt
 F   G   E   L   C   H   L   A   L   -   H   L   C   L   S   H   W   P   P   L
ctt gtg ggc act ttc tgg atg ccc agg cct aat cca gcg ctg gca ctc agt taa ttc tct
 L   V   G   T   F   W   M   P   R   P   N   P   A   L   A   L   S   -   F   S
ctt cct tta aag gga aca att acc acc ttg agg aag gag gcc tga aag cta agt ctt tgc
 L   P   L   K   G   T   I   T   T   L   R   K   E   A   -   K   L   S   L   C
tat tgc atc cgc ctt ctt ctc acg aag acc tgc agc ttg aaa gga ggg gag gga gaa ata
 Y   C   I   R   L   L   L   T   K   T   C   S   L   K   G   G   E   G   E   I
gcc cta agc tcc agg aag acc aag
 A   L   S   S   R   K   T   K
```

2) **LncRNA ENST00000811416.1**

rs61871745

**5'3' Frame 1**

```
gct ggt ggc ttc atc agg aag gga gtc agc agg ggg agg act ggg gga cca agc ggt ccc
 A   G   G   F   I   R   K   G   V   S   R   G   R   T   G   G   P   S   G   P
taa gaa gca gaa ttt gca ata ccc aag gtt tta gaa aga gca acc tgg cag ggc cac tgc
 -   E   A   E   F   A   I   P   K   V   L   E   R   A   T   W   Q   G   H   C
aca atg aga tca cag act cac ctc cca gct gtg agt gca gct cac ccc acc tgc tct gac
 T   M   R   S   Q   T   H   L   P   A   V   S   A   A   H   P   T   C   S   D
cca ggc acc ggt tat gaa gaa gac aag cag ctc tac aga ggg gcc tca aga aat cct tcg
 P   G   T   G   Y   E   E   D   K   Q   L   Y   R   G   A   S   R   N   P   S
aaa cag aaa tca aag tct tgc tgg aag ggt tct ggg cac cct gga ctg tcc aca tgg gag
 K   Q   K   S   K   S   C   W   K   G   S   G   H   P   G   L   S   T   W   E
gct gtt ggt gcg tgt gcc tac gtg tgt cca tca gtg gat gca cga aga agg aaa tga att
 A   V   G   A   C   A   Y   V   C   P   S   V   D   A   R   R   R   K   -   I
gat tcc att ctt gat gtg act ttc agc tgt cag ctg tct gca ctc cca gac ggg gtg atg
 D   S   I   L   D   V   T   F   S   C   Q   L   S   A   L   P   D   G   V   M
gcc tgg cac caa cgg agc att acc ctg atg aat cat ggt cga ggg aac aat tac cac ctt  G/A=C/T, Thr to Ile
 A   W   H   Q   R   S   I   T   L   M   N   H   G   R   G   N   N   Y   H   L
gag gaa gga ggc ctg aaa gct aag tct ttg cta ttg cat ccg cct tct tct cac gaa gac
 E   E   G   G   L   K   A   K   S   L   L   L   H   P   P   S   S   H   E   D
ctg cag ctt gaa agg agg gga ggg aga aat agc cct aag ctc cag gaa gac caa gcc cag
 L   Q   L   E   R   R   G   G   R   N   S   P   K   L   Q   E   D   Q   A   Q
cct ctg tga tga ttc cga gga aat tca gga aat cct tgc taa tgg cca aac ttc ccg ggt
 P   L   -   -   F   R   G   N   S   G   N   P   C   -   W   P   N   F   P   G
ttg aga tgc aaa agg aaa tca gat cgc agg gta atg ctt tta cac taa gca aga aat tgc
 L   R   C   K   R   K   S   D   R   R   V   M   L   L   H   -   A   R   N   C
agg ata taa agc tgt gca gac cat tgc ctc tga tta aga aa
 R   I   -   S   C   A   D   H   C   L   -   L   R
```

**5'3' Frame 2**

```
gctg gtg gct tca tca gga agg gag tca gca ggg gga gga ctg ggg gac caa gcg gtc cct
  L   V   A   S   S   G   R   E   S   A   G   G   G   L   G   D   Q   A   V   P
aag aag cag aat ttg caa tac cca agg ttt tag aaa gag caa cct ggc agg gcc act gca
 K   K   Q   N   L   Q   Y   P   R   F   -   K   E   Q   P   G   R   A   T   A
caa tga gat cac aga ctc acc tcc cag ctg tga gtg cag ctc acc cca cct gct ctg acc
 Q   -   D   H   R   L   T   S   Q   L   -   V   Q   L   T   P   P   A   L   T
cag gca ccg gtt atg aag aag aca agc agc tct aca gag ggg cct caa gaa atc ctt cga
 Q   A   P   V   M   K   K   T   S   S   S   T   E   G   P   Q   E   I   L   R
aac aga aat caa agt ctt gct gga agg gtt ctg ggc acc ctg gac tgt cca cat ggg agg
 N   R   N   Q   S   L   A   G   R   V   L   G   T   L   D   C   P   H   G   R
ctg ttg gtg cgt gtg cct acg tgt gtc cat cag tgg atg cac gaa gaa gga aat gaa ttg
 L   L   V   R   V   P   T   C   V   H   Q   W   M   H   E   E   G   N   E   L
att cca ttc ttg atg tga ctt tca gct gtc agc tgt ctg cac tcc cag acg ggg tga tgg
 I   P   F   L   M   -   L   S   A   V   S   C   L   H   S   Q   T   G   -   W
cct ggc acc aac gga gca tta ccc tga tga atc atg gtc gag gga aca att acc acc ttg
 P   G   T   N   G   A   L   P   -   -   I   M   V   E   G   T   I   T   T   L
agg aag gag gcc tga aag cta agt ctt tgc tat tgc atc cgc ctt ctt ctc acg aag acc
 R   K   E   A   -   K   L   S   L   C   Y   C   I   R   L   L   L   T   K   T
tgc agc ttg aaa gga ggg gag gga gaa ata gcc cta agc tcc agg aag acc aag ccc agc
 C   S   L   K   G   G   E   G   E   I   A   L   S   S   R   K   T   K   P   S
ctc tgt gat gat tcc gag gaa att cag gaa atc ctt gct aat ggc caa act tcc cgg gtt
 L   C   D   D   S   E   E   I   Q   E   I   L   A   N   G   Q   T   S   R   V
tga gat gca aaa gga aat cag atc gca ggg taa tgc ttt tac act aag caa gaa att gca
 -   D   A   K   G   N   Q   I   A   G   -   C   F   Y   T   K   Q   E   I   A
gga tat aaa gct gtg cag acc att gcc tct gat taa gaa a
 G   Y   K   A   V   Q   T   I   A   S   D   -   E
```

**5'3' Frame 3**

```
gctgg tgg ctt cat cag gaa ggg agt cag cag ggg gag gac tgg ggg acc aag cgg tcc cta
   W   W   L   H   Q   E   G   S   Q   Q   G   E   D   W   G   T   K   R   S   L
aga agc aga att tgc aat acc caa ggt ttt aga aag agc aac ctg gca ggg cca ctg cac
 R   S   R   I   C   N   T   Q   G   F   R   K   S   N   L   A   G   P   L   H
aat gag atc aca gac tca cct ccc agc tgt gag tgc agc tca ccc cac ctg ctc tga ccc
 N   E   I   T   D   S   P   P   S   C   E   C   S   S   P   H   L   L   -   P
agg cac cgg tta tga aga aga caa gca gct cta cag agg ggc ctc aag aaa tcc ttc gaa
 R   H   R   L   -   R   R   Q   A   A   L   Q   R   G   L   K   K   S   F   E
aca gaa atc aaa gtc ttg ctg gaa ggg ttc tgg gca ccc tgg act gtc cac atg gga ggc
 T   E   I   K   V   L   L   E   G   F   W   A   P   W   T   V   H   M   G   G
tgt tgg tgc gtg tgc cta cgt gtg tcc atc agt gga tgc acg aag aag gaa atg aat tga
 C   W   C   V   C   L   R   V   S   I   S   G   C   T   K   K   E   M   N   -
ttc cat tct tga tgt gac ttt cag ctg tca gct gtc tgc act ccc aga cgg ggt gat ggc
 F   H   S   -   C   D   F   Q   L   S   A   V   C   T   P   R   R   G   D   G
ctg gca cca acg gag cat tac cct gat gaa tca tgg tcg agg gaa caa tta cca cct tga
 L   A   P   T   E   H   Y   P   D   E   S   W   S   R   E   Q   L   P   P   -
gga agg agg cct gaa agc taa gtc ttt gct att gca tcc gcc ttc ttc tca cga aga cct
 G   R   R   P   E   S   -   V   F   A   I   A   S   A   F   F   S   R   R   P
gca gct tga aag gag ggg agg gag aaa tag ccc taa gct cca gga aga cca agc cca gcc
 A   A   -   K   E   G   R   E   K   -   P   -   A   P   G   R   P   S   P   A
tct gtg atg att ccg agg aaa ttc agg aaa tcc ttg cta atg gcc aaa ctt ccc ggg ttt
 S   V   M   I   P   R   K   F   R   K   S   L   L   M   A   K   L   P   G   F
gag atg caa aag gaa atc aga tcg cag ggt aat gct ttt aca cta agc aag aaa ttg cag
 E   M   Q   K   E   I   R   S   Q   G   N   A   F   T   L   S   K   K   L   Q
gat ata aag ctg tgc aga cca ttg cct ctg att aag aaa
 D   I   K   L   C   R   P   L   P   L   I   K   K
```

3. **Table S2 HTRA1-AS1 Variants Information**

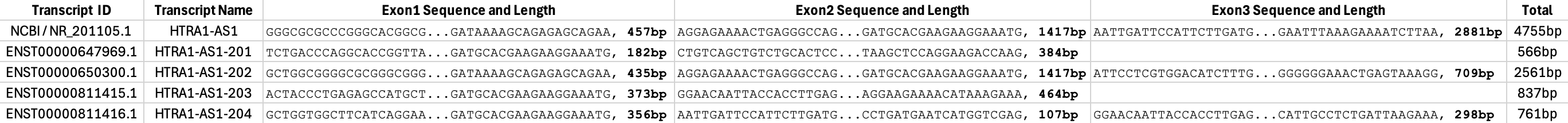

4. **Table S2 RNA Integrity Metrics (RIM) of Human Retinal RNA Samples from Control and AMD Donors**

| **Human Retinal RNA** | **Mean of RIM** | **SD of RIM** | **Samples size (n)** |
| --- | --- | --- | --- |
| Control (CTRL) | 0.814 | 0.056 | 44 |
| Age-related macular degeneration (AMD) | 0.823 | 0.051 | 43 |

**5. Table S4. AMD associated common SNPs in the HTRA1AS1/ARMS2 gene region**

|  | **SNP** | **GRCh38 position** | **Alleles** |
| --- | --- | --- | --- |
|  | rs61871745 | 10:122450853 | G/A |
|  | rs11200632 | 10:122452020 | A/G |
|  | rs11200633 | 10:122452080 | C/T |
|  | rs7898343 | 10:122453371 | G/C |
|  | rs61871746 | 10:122453397 | T/C |
|  | rs6585831 | 10:122453429 | T/C |
|  | rs61871747 | 10:122453530 | C/T |
|  | rs370974631 | 10:122453628 | AA/- |
|  | rs2672600 | 10:122453723 | G/C |
|  | rs72631113 | 10:122453933 | C/T |
|  | rs200227426 | 10:122454155 | C/A |
|  | rs370728615 | 10:122454155 | CAACAACAAC/- |
|  | rs201396317 | 10:122454158 | C/A |
|  | rs199637836 | 10:122454161 | C/A |
|  | rs11200634 | 10:122454164 | C/A |
|  | rs142585532 | 10:122454173 | -/AAAAAAAAAAAAAA |
|  | rs2736911 | 10:122454839 | C/T |
|  | rs10490924 | 10:122454932 | G/T |
|  | rs61544945 | 10:122455085 | -/GT |
|  | rs36212731 | 10:122455460 | G/T |
|  | rs36212732 | 10:122455682 | A/G |
|  | rs36212733 | 10:122455695 | T/C |
|  | rs3750848 | 10:122455799 | T/G |
|  | rs3750847 | 10:122455905 | C/T |
|  | rs3750846 | 10:122456049 | T/C |

Note: Only AMD-associated SNP rs61871745 (G/A; A as the risk allele) is located within predicted coding regions of *HTRA1-AS1* and *ENST00000647969.1 (*highlighted*).* The risk allele A may induce nucleotide substitution (C→T) that changes the predicted encoded codon from ACC to ATC (Thr→Ile) within one of the putative open reading frames.
